# Age could be driving variable SARS-CoV-2 epidemic trajectories worldwide

**DOI:** 10.1101/2020.04.13.20059253

**Authors:** Houssein H. Ayoub, Hiam Chemaitelly, Shaheen Seedat, Ghina R. Mumtaz, Monia Makhoul, Laith J. Abu-Raddad

## Abstract

**Background:** Current geographic spread of documented severe acute respiratory syndrome coronavirus 2 (SARS-CoV-2) infections shows heterogeneity. This study explores the role of age in potentially driving differentials in infection spread, epidemic potential, and rates of disease severity and mortality across countries.

**Methods:** An age-stratified deterministic mathematical model that describes SARS-CoV-2 transmission dynamics was applied to 159 countries and territories with a population ≥1 million.

**Results:** Assuming worst-case scenario for the pandemic, the results indicate that there could be stark regional differences in epidemic trajectories driven by differences in the distribution of the population by age. In the African Region (median age: 18.9 years), the median *R*_0_ was 1.05 versus 2.05 in the European Region (median age: 41.7 years), and the median (per 100 persons) for the infections rate was 22.5 (versus 69.0), for severe and/or critical disease cases rate was 3.3 (versus 13.0), and for death rate was 0.5 (versus 3.9).

**Conclusions:** Age could be a driver of variable SARS-CoV-2 epidemic trajectories worldwide. Countries with sizable adult and/or elderly populations and smaller children populations may experience large and rapid epidemics in absence of interventions. Meanwhile, countries with predominantly younger age cohorts may experience smaller and slower epidemics. These predictions, however, should not lead to complacency, as the pandemic could still have a heavy toll nearly everywhere.

## INTRODUCTION

The current geographic spread of the documented severe acute respiratory syndrome coronavirus 2 (SARS-CoV-2) [1] infections and associated Coronavirus Disease 2019 (COVID-2019) [1] shows heterogeneity [2]. This remains unexplained but may possibly reflect delays in virus introduction into the population, differentials in testing and/or reporting, differentials in the implementation, scale, adherence, and timing of public health interventions, or other epidemiological factors. In this study, we explore the role of age in explaining the differential spread of the infection and its future epidemic potential.

As our understanding of the SARS-CoV-2 transmission dynamics is rapidly evolving [3], the role of age in the epidemiology of this infection is becoming increasingly apparent [4-7]. In a recent study examining SARS-CoV-2 epidemiology in China [4], we quantified the effect of age on the *biological* susceptibility to infection acquisition. We found that *relative* to individuals aged 60-69 years, susceptibility was 0.06 in those aged 0-19 years, 0.34 in those aged 20-29 years, 0.57 in those aged 30-39 years, 0.69 in those aged 40-49 years, 0.79 in those aged 50-59 years, 0.94 in those aged 70-79 years, and 0.88 in those aged ≥80 years (Supplementary Figure S1). Notably, this age-dependence was estimated after accounting for the assortativeness in population mixing by age [4].

Age also affects disease progression [7-10] and mortality risk [11-13] among those infected. The proportion of infections that eventually progress to severe disease, critical disease, or death, increases rapidly with age, especially among those ≥50 years of age (Supplementary Figure S2) [8-12]. Since the demographic structure of the population (that is the distribution of the population across the different age groups) varies by country and region, this poses a question as to the extent to which age effects can drive geographic differentials in the reproduction number (*R*_0_), epidemic potential, and rates of disease severity and mortality.

We aimed here to answer this question and to estimate for each country (with a population ≥1 million), region, and globally, *R*_0_, and the rate per 100 persons (out of the total population by the end of the epidemic cycle) of each of the cumulative number of incident infections, mild infections, severe and/or critical disease cases, and deaths, in addition to the number of days needed for the national epidemic to reach its incidence peak (a measure of how fast the epidemic will grow). We also aimed to introduce indices for infection severity, mortality, and susceptibility across countries.

## METHODS

We adapted and applied our recently developed deterministic mathematical model [4] describing SARS-CoV-2 transmission dynamics in China (Supplementary Figure S3), to 159 countries and territories, virtually covering the world population [14]. Since our focus was on investigating the natural course of the SARS-CoV-2 epidemic and on assessing its full epidemic potential, the model was applied assuming the *worst-case* scenario for the epidemic in each country, that is in absence of any intervention.

The model stratified the population into compartments according to age (0-9, 10-19, …, ≥80 years), infection status (uninfected, infected), infection stage (mild, severe, critical), and disease stage (severe, critical), using a system of coupled nonlinear differential equations (Supplementary Section 1). Susceptible individuals in each age group were assumed at risk of acquiring the infection at a hazard rate that varies based on the age-specific susceptibility to the infection (Supplementary Figure S1), the infectious contact rate per day, and the transmission mixing matrix between the different age groups (Supplementary Section 1). Following a latency period of 3.69 days [15-18], infected individuals develop mild, severe, or critical infection, as informed by the observed age-specific distribution of cases across these infection stages in China [8-10]. The duration of infectiousness was assumed to last for 3.48 days [8, 15, 17, 18] after which individuals with mild infection recover, while those with severe and critical infection develop, respectively, severe and critical disease over a period of 28 days [8] prior to recovery. Individuals with critical disease have the additional risk of disease mortality, as informed by the age-stratified disease mortality rate in China [11, 12].

Model parameters were based on current data for SARS-CoV-2 natural history and epidemiology, or through model fitting to the China outbreak data [4] (Supplementary Section 2 and Table S1). Namely, the overall infectious contact rate, age-specific biological susceptibility profile, and age-specific mortality rate were assumed as those in China [4]. The population size, demographic structure, and life expectancy in countries and territories with a population of ≥1 million, as of 2020, were extracted from the United Nations World Population Prospects database [14]. Model parameters, definitions, values, and justifications are in Supplementary Section 2 and Tables S1 and S2. Indices for mortality, infection severity, and susceptibility were derived by weighting each of these factors in each age group by the fraction of the population in that age group, and then summing the contributions of all ages.

Ranges of uncertainty around model-predicted outcomes were determined using five-hundred simulation runs that applied Latin Hypercube sampling [19, 20] from a multidimensional distribution of the model parameters. At each run, input parameter values were selected from ranges specified by assuming ±30% uncertainty around parameters’ point estimates. The resulting distribution for each model-predicted outcome was then used to derive the most probable estimate and 95% uncertainty interval (UI). Mathematical modelling analyses were performed in MATLAB R2019a [21], while statistical analyses were performed in STATA/SE 16.1 [22].

## RESULTS

In what follows, we highlight results for select (mostly populous) countries that are of broad geographic representation and characterized by diverse demographic structures. These include Brazil, China, Egypt, India, Indonesia, Italy, Niger, Pakistan, and USA. Detailed results for all 159 countries and territories are in Supplementary Tables S3-S8.

Figure 1A shows the estimated *R*_0_ in these select countries, which was highest in Italy at 2.30, followed by USA and China at 1.95, Brazil at 1.75, Indonesia at 1.55, India at 1.45, Egypt at 1.35, Pakistan at 1.25, and lowest in Niger at 0.93. Country-specific estimates of *R*_0_ grouped by World Health Organization (WHO) region are illustrated in Figure 2. The median *R*_0_ was 1.05 (range: 0.93-1.95) in African Region (AFRO), 1.45 (range: 0.98-1.75) in Eastern Mediterranean Region (EMRO), 1.55 (range: 1.15-1.95) in South-East Asia Region (SEARO), 1.65 (range: 1.25-2.28) in Region of the Americas (AMRO), 1.85 (range: 1.25-2.30) in Western pacific Region (WPRO), and 2.05 (range: 1.25-2.30) in European Region (EURO). Globally, the median *R*_0_ was 1.55 with a range of 0.93-2.30.

**Figure 1:**
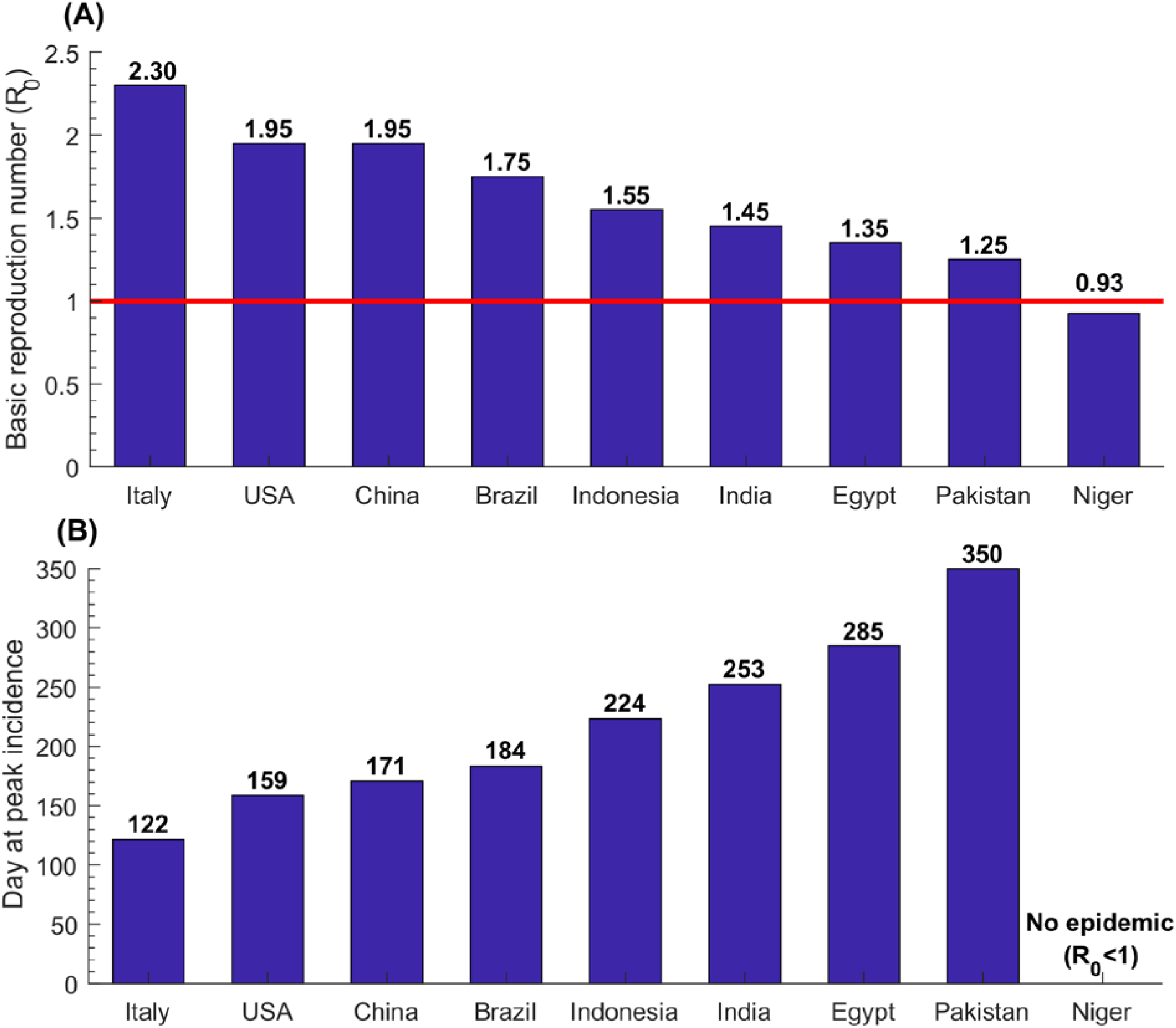
**Estimates for the basic reproduction number, *R*_0_, and the number of days needed for the national epidemic to reach its incidence peak, in select countries.**

**Figure 2:**
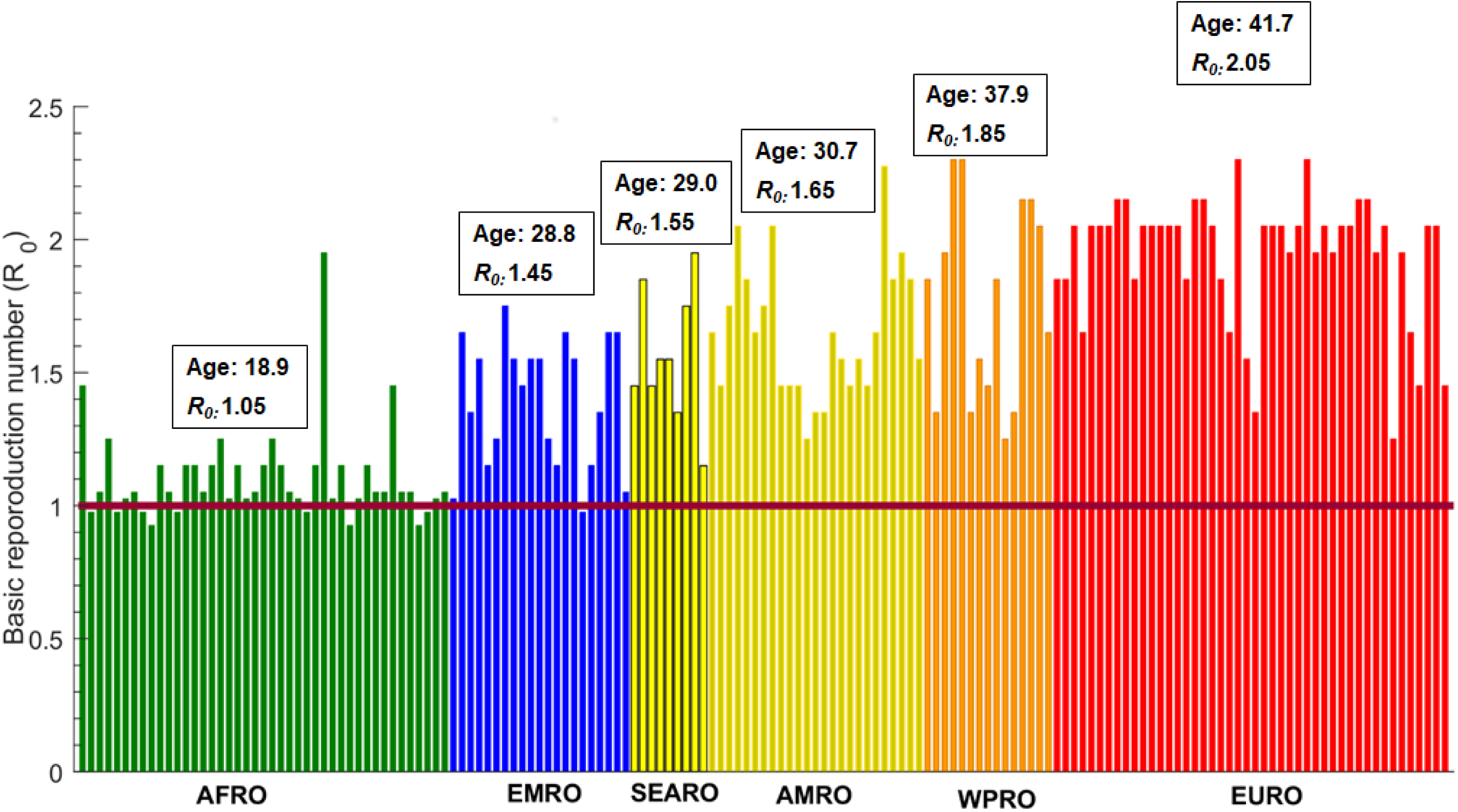
Estimates for the basic reproduction number *R*_0_ in 159 countries and territories with a population of at least one million, across World Health Organization regions. These are the African Region (AFRO), Eastern Mediterranean Region (EMRO), South-East Asia Region (SEARO), Region of the Americas (AMRO), Western Pacific Region (WPRO), and European Region (EURO). The figure shows also the median age in years and the median *R*_0_ for each world region.

Figure 1B shows the estimated number of days needed for the national epidemic to reach its incidence peak. In Italy, the peak was reached after 122 days (∼4 months), whereas in Pakistan it was reached after 350 days (nearly a year). Meanwhile, since *R*_0_ <1, no epidemic would emerge in Niger. Of note that the number of days needed for the national epidemic to reach its incidence peak depends on the population size in addition to *R*_0_ — it takes more time for the epidemic to reach its peak in larger nations (Supplementary Tables S3-S8).

The estimated incidence rate per 100 persons, defined as the *cumulative* number of infections by the end of the epidemic cycle out of the total population, was highest in Italy at 75.5 and lowest in Pakistan at 33.0 (Figure 3A). Across world regions, the median incidence rate per 100 persons was 22.5 (range: 0.25-63.5) in AFRO, 45.0 (range: 1.5-65.0) in EMRO, 49.0 (range: 25.0-66.5) in SEARO, 53.0 in AMRO (range: 35.0-70.5), 63.5 (range: 31.0-76.5) in WPRO, and 69.0 (range: 31.0-75.5) in EURO, and 49.0 (range: 0.25-76.5) globally.

**Figure 3:**
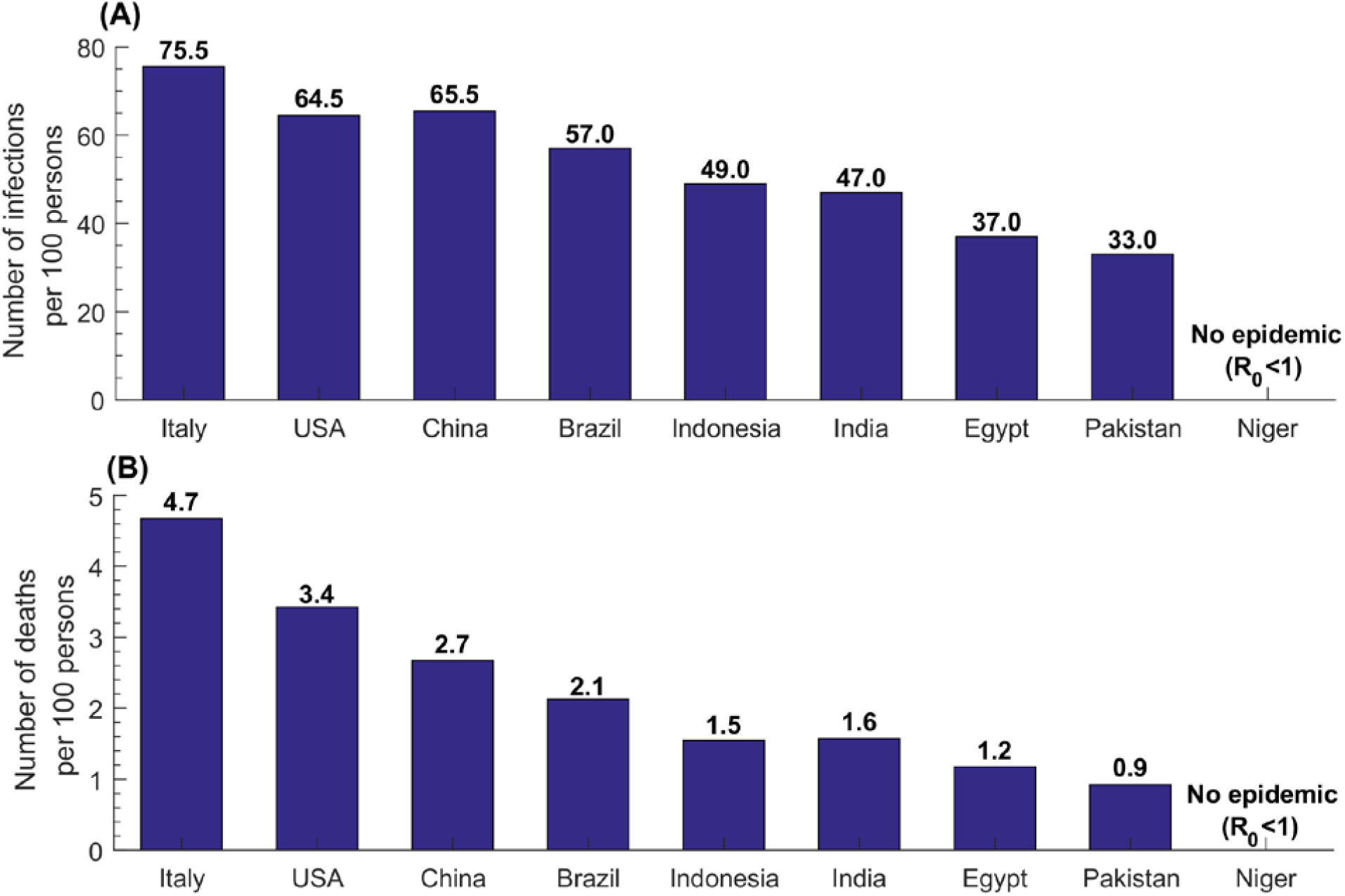
**Estimates for the number of infections and the number of deaths per 100 persons in select countries.**

The estimated death rate per 100 persons followed the same pattern, where it was highest in Italy at 4.7 and lowest in Pakistan at 0.9 (Figure 3B). Across world regions, the median death rate per 100 persons was 0.5 (range: 0.0-2.8) in AFRO, 0.9 (range: 0.0-2.0) in EMRO, 1.6 (range: 0.8- 2.9) in SEARO, 2.0 in AMRO (range: 1.0-4.2), 2.7 (range: 0.8-5.3) in WPRO, and 3.9 (range: 0.8-4.7) in EURO, and 1.6 (range: 0.0-5.3) globally.

The estimated rate of mild infections per 100 persons was highest in Italy at 60.5 and lowest in Pakistan at 27.0 (Figure 4A). Across world regions, the median per 100 persons was 17.0 (range: 0.25-52.5) in AFRO, 39.0 (range: 1.5-55.0) in EMRO, 41.0 (range: 23.0-54.5) in SEARO, 43.5 in AMRO (range: 29.0-56.5), 51.5 (range: 27.0-61.5) in WPRO, and 56.5 (range: 25.0-65.0) in EURO, and 41.0 (range: 0.25-61.5) globally.

**Figure 4:**
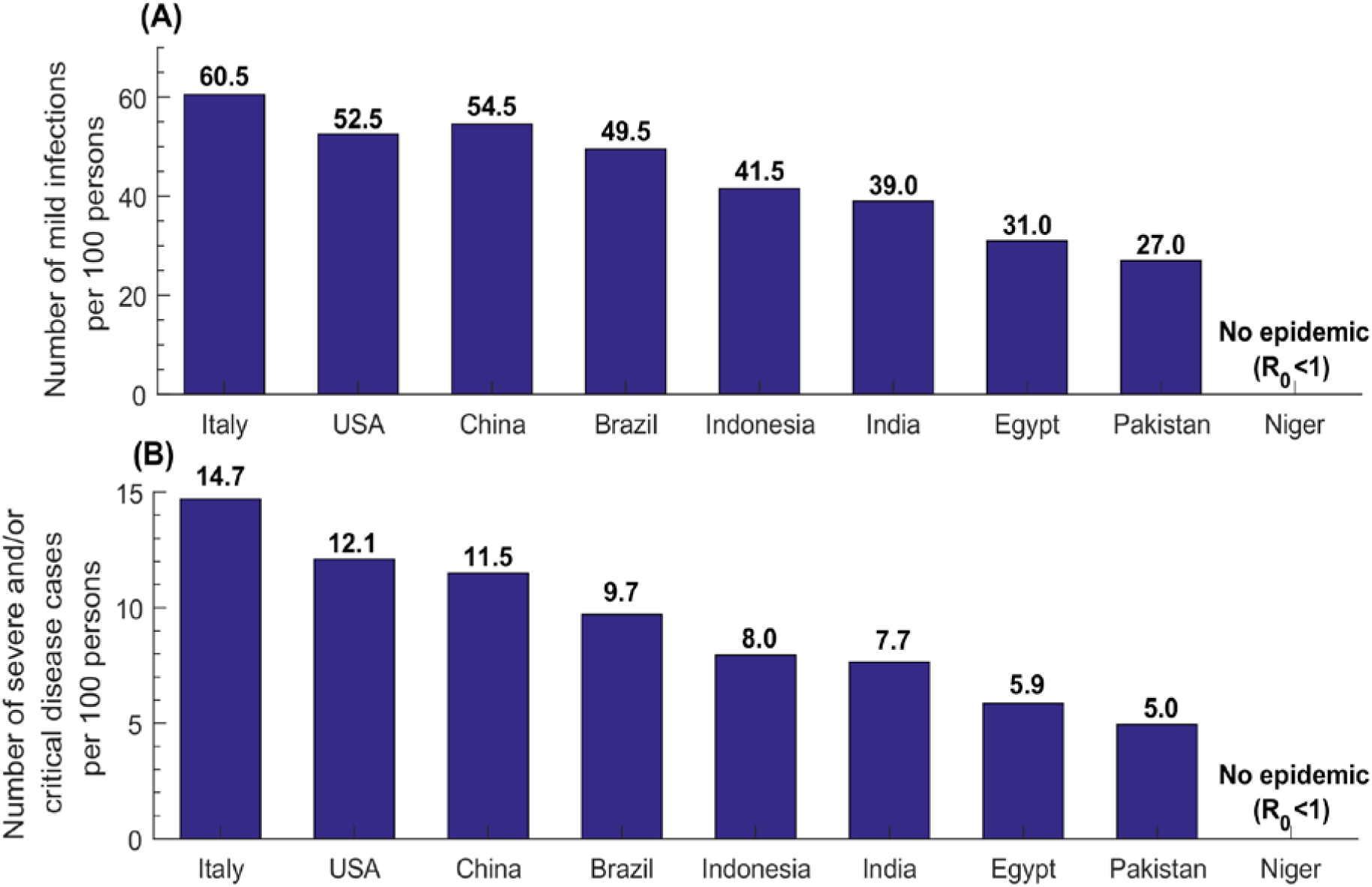
**Estimates for the number of mild infections and the number of severe and/or critical diseases cases per 100 persons in select countries.**

Meanwhile, the estimated rate of severe and/or critical disease cases per 100 persons was highest in Italy at 14.7 and lowest in Pakistan at 5.0 (Figure 4B). Across world regions, the median per 100 persons was 3.3 (range: 0.0-11.3) in AFRO, 7.1 (range: 0.3-9.5) in EMRO, 7.7 (range: 4.1- 11.9) in SEARO, 8.7 in AMRO (range: 5.0-13.5), 11.5 (range: 5.0-15.5) in WPRO, and 13.0 (range: 4.7-14.7) in EURO, and 8.0 (range: 0.0-15.5) globally.

The indices for mortality, infection severity, and susceptibility varied across countries and regions (Figure 5 and Supplementary Figure S4). The medians for the mortality, critical infections, and susceptibility indices were lowest in AFRO at 0.5%, 2.7%, and 28.2%, respectively, and highest in EURO at 2.4%, 7.4%, and 53.7%, respectively (Figure 5). Meanwhile, the medians for mild infections and severe infections indices were lowest in EURO at 83.1% and 9.6%, respectively and highest in AFRO at 87.2% and 10.2%, respectively (Supplementary Figure S4).

**Figure 5:**
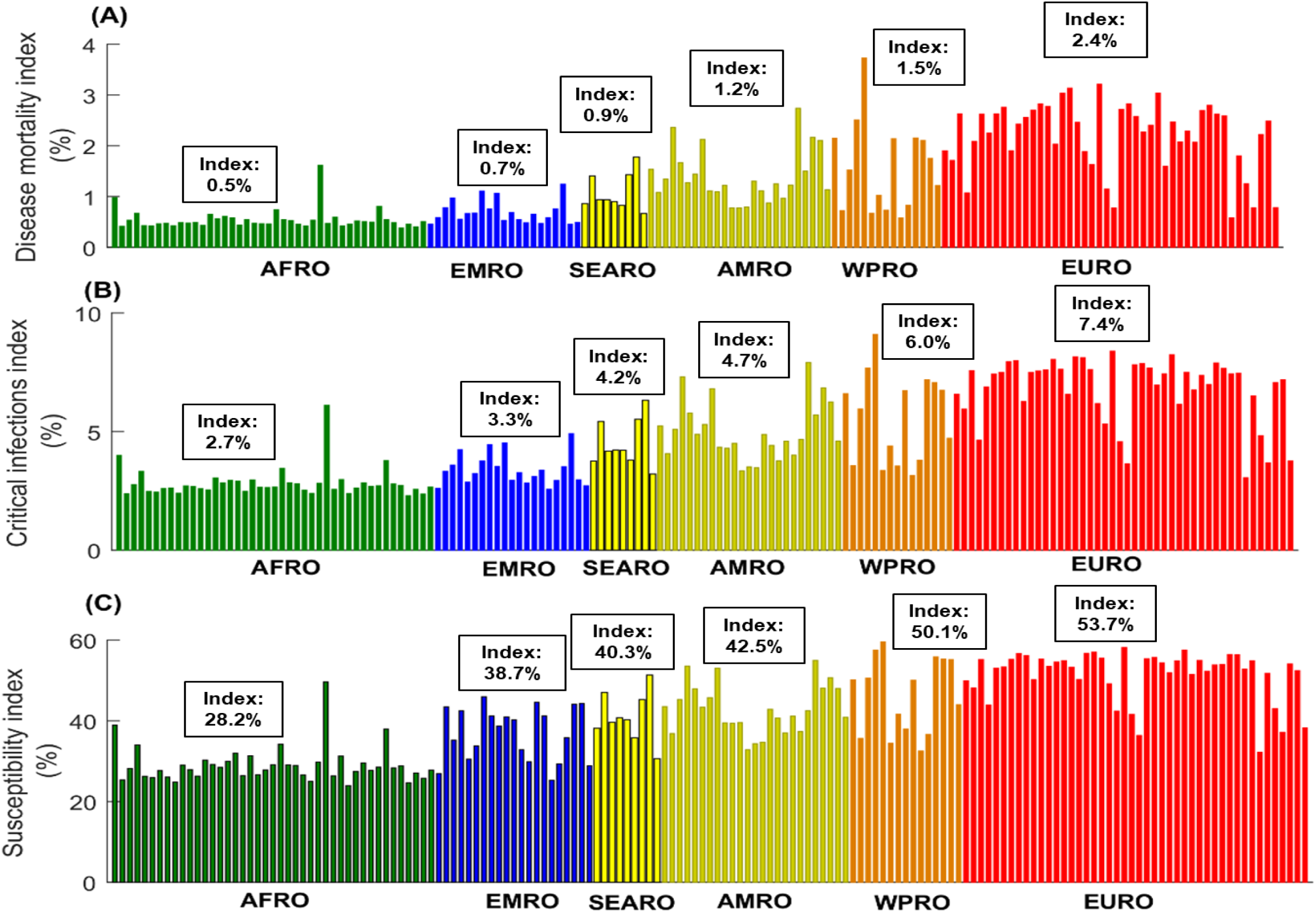
Estimates for the A) disease mortality index, B) critical infections index, and C) susceptibility index in 159 countries and territories with a population of at least one million, across World Health Organization regions. These are the African Region (AFRO), Eastern Mediterranean Region (EMRO), South-East Asia Region (SEARO), Region of the Americas (AMRO), Western Pacific Region (WPRO), and European Region (EURO). The figure shows also the median index for each world region.

## DISCUSSION

The above results suggest that although this pandemic is a formidable challenge globally, its intensity and toll in terms of morbidity and mortality may vary substantially by country and regionally. While some countries, particularly in resource-rich countries, may experience large and rapid epidemics, other countries, particularly in resource-poor countries, may experience smaller and slower epidemics. While the scale of the epidemic may be smallest in sub-Saharan Africa with its young population, it could be intense in countries with small children population and sizable adult and/or elderly population.

The key finding of this study is the role of age in potentially driving divergent SARS-CoV-2 epidemic trajectories and disease burdens. The effect of age reflects the interplay of three main factors: the variation in susceptibility by age (Supplementary Figure S1), variation in disease severity and mortality by age (Supplementary Figure S2), and variation in demographic structure across countries (Supplementary Tables S3-S8). The combined effect of these factors can result in stark variations in *R*_0_ (Figure 2), and consequently epidemic potential, disease severity, and disease mortality—there was strong correlation between *R*_0_ and median age across countries (Supplementary Figure S5). Incidentally, the globally diagnosed cases as of April 9^th^, 2020 show a correlation with median age across countries (Spearman correlation coefficient: 0.71 (95% CI: 0.62-0.78), p<0.001). A linear regression analysis using these data indicated an increase in the number of diagnosed infections by 1058.0 (95% CI: 386.0-1729.9; p: 0.002) for every one-year increase in median age. Although this may be explained by most diagnosed infections being reported by countries with advanced testing infrastructure, this may support the role of age presented here. Importantly, as infection transmission dynamics is a non-linear phenomenon, even small changes in *R*_0_, when *R*_0_ is in the range of 1-2, can drive considerable differences in epidemic size and trajectory (Supplementary Figure S6).

An illustration of the role of age can be seen in Niger, a nation with a median age of only 15 years, where the exclusion of children from the population would have increased *R*_0_ from 0.93 to 2.6. This demonstrates how the lower susceptibility among younger persons, particularly children, acts as a “herd immunity” impeding the ferocious strength of the force of infection of an otherwise very infectious virus. This epidemiological feature contrasts that of other respiratory infections, such as the 2009 influenza A (H1N1) pandemic (H1N1pdm) infection (Supplementary Figure S7) [23], where the cumulative incidence was found to be highest among children and young adults, and much smaller among older adults.

One possible inference to be drawn from the above results is that epidemic size and associated disease burden could be highest in settings with sizable mid-age and/or elderly populations, as currently exemplified by Italy. This may also possibly explain sub-national patterns, such as the large number of diagnosed cases in the city of New York, although additional fine-grained analyses are needed to delineate within-country heterogeneities in transmission dynamics.

Another possible inference of relevance to containment efforts relates to the likelihood of the infection establishing itself in a population. The probability of a major outbreak upon introduction of one infection is given approximately by 1−1 *R*_0_ [24, 25]. Accordingly, in countries where *R*_0_ is just above the epidemic threshold of *R*_0_ = 1, the virus will need to be introduced multiple times before it can generate sustainable chains of transmission (Supplementary Figure S8 provides an illustration of this effect). In such countries, less disruptive social distancing measures may be sufficient to contain the epidemic compared to countries with larger *R*_0_.

Our study explores the potential effect of age on the epidemiology of SARS-CoV-2, but other factors that remain poorly understood, may also contribute to driving different epidemic trajectories. Transmission of the virus may be affected by seasonality, environmental and genetic factors, differences in social network structure and cultural norms (such as shaking hands, kissing, and other person-to-person contacts), and underlying co-morbidities which also impact disease severity and mortality. These factors may have contributed to a slowly growing epidemic in Japan, despite its demographic structure, as opposed to fast growing epidemics in the European Region and the USA.

This study has limitations. Model estimations are contingent on the validity and generalizability of input data. Our estimates were based on SARS-CoV-2 natural history and disease progression data from China, but these may not be applicable to other countries. The key factor driving the heterogeneity in transmission dynamics is the age-specific biological susceptibility profile. We used the susceptibility profile as derived from the China outbreak data [4], but this may not be generalizable to other countries. More estimates from other countries are needed to investigate the potential variation in this profile. While current data on the attack rate from different countries supports age heterogeneity and lower exposure among children, different countries show still variation in the age-stratified attack rate (Supplementary Figures S9 and S10). Of note that the observed lower susceptibility to the infection at younger age [4] does not necessarily imply absence of infection, but may reflect rapid infection clearance or subclinical infection. This may impact our estimates depending on these infections’ degree of infectiousness. A sensitivity analysis assuming a 50% increase in the susceptibility of younger age cohorts (to somewhat reflect the effect of subclinical infections) resulted in smaller differences in *R*_0_ across countries, but still supported our findings of wide heterogeneity in *R*_0_ (supplementary Figure S11). It is conceivable that such subclinical infections may have lower viral load, and therefore rapid infection clearance and limited transmission potential. Data on infection clusters from China and other countries suggested that children do not appear to play a significant role in the transmission of this infection [7, 8, 26].

Our study may have overestimated disease mortality by basing mortality rates on estimates of crude case fatality rate in China, as suggested by a recent study [13]. Our results are based on the most probable value for *R*_0_ as estimated through 500 runs of uncertainty analysis. The latter may have biased our reported results towards lower *R*_0_. For instance, the most probable value for *R*_0_ in China was 1.95, but the point estimate assuming the baseline values of the input parameters was higher at 2.10 [4]. Despite these limitations, our parsimonious model, tailored to the nature of available data, was able to reproduce the epidemic as observed in China [4], and generated results that are valid to a wide range of model assumptions.

## CONCLUSION

Age appears to be a driver of variable SARS-CoV-2 epidemic trajectories worldwide. Countries with sizable adult and/or elderly populations and smaller children populations may experience large and rapid epidemics in absence of interventions, necessitating disruptive social distancing measures to contain the epidemic. Meanwhile, countries with predominantly younger age cohorts may experience smaller and slower epidemics and may require less disruptive social distancing measures to contain the epidemic. These predictions, however, should not lead to complacency, and should not affect national response in strengthening preparedness plans and in implementing prevention interventions. Once established, and even if smaller in scale due to their predominantly younger population, the SARS-CoV-2 epidemic would still cause a heavy toll on developing countries with resource-poor healthcare infrastructure.

## Data Availability

All data generated or analysed during this study are included in this article and its Supplementary Information file.

## Acknowledgements

This publication was made possible by NPRP grant number 9-040-3-008 and NPRP grant number 12S-0216-190094 from the Qatar National Research Fund (a member of Qatar Foundation). GM acknowledges support by UK Research and Innovation as part of the Global Challenges Research Fund, grant number ES/P010873/1. The statements made herein are solely the responsibility of the authors. The authors are also grateful for support provided by the Biomedical Research Program and the Biostatistics, Epidemiology, and Biomathematics Research Core, both at Weill Cornell Medicine-Qatar.

## Author contributions

HHA constructed and parameterized the mathematical model and conducted the mathematical modelling analyses. HC contributed to the parameterization of the mathematical model, conducted the statistical analyses, and wrote the first draft of the manuscript. LJA conceived and led the design of the study, construct and parameterization of the mathematical model, conduct of analyses, and drafting of the article. All authors contributed to discussion and interpretation of the results and to the writing of the manuscript. All authors have read and approved the final manuscript.

---

**Figure S1:**
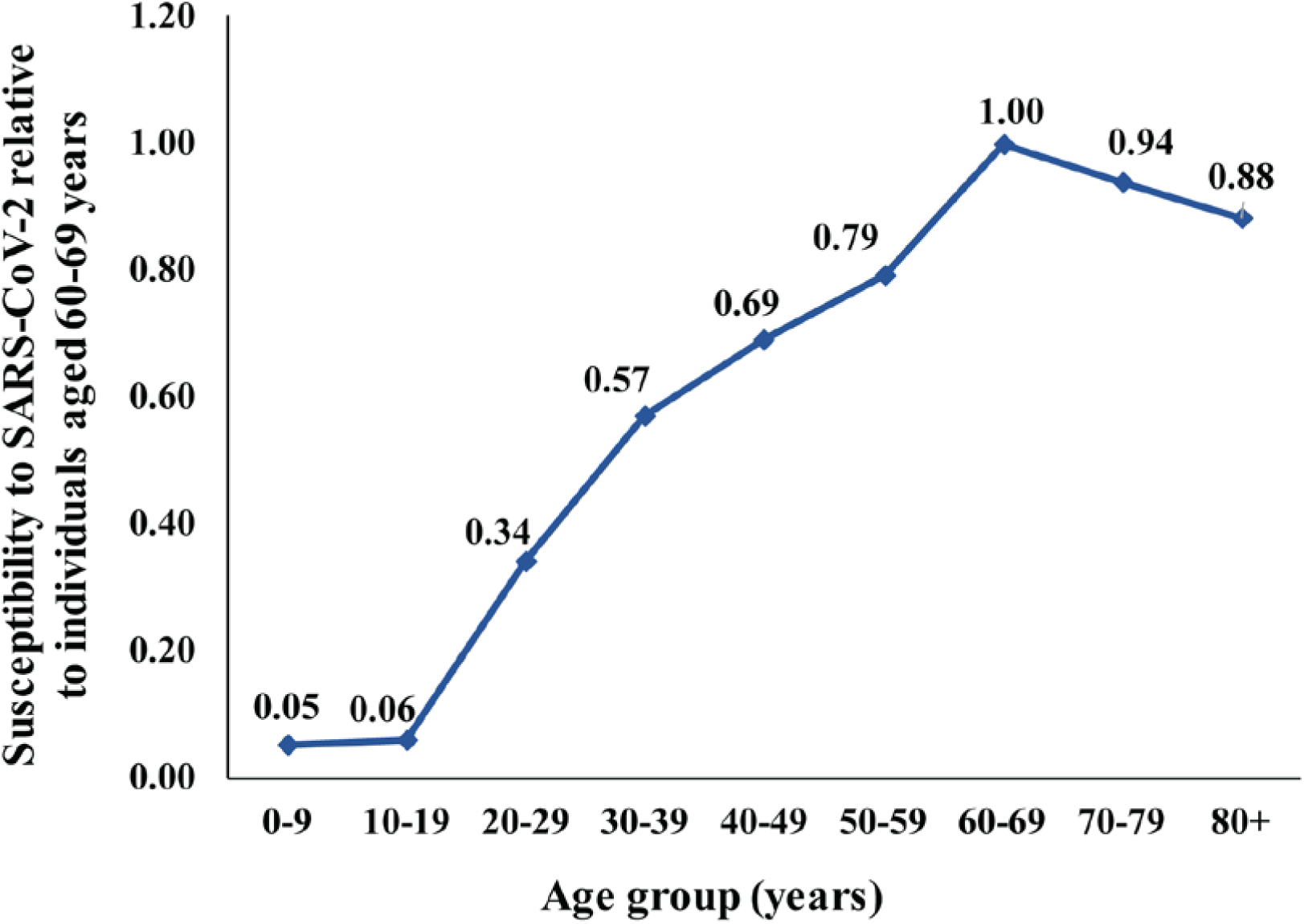
**Age-specific susceptibility to SARS-CoV-2 acquisition relative to individuals aged 60-69 years [1].**

**Figure S2:**
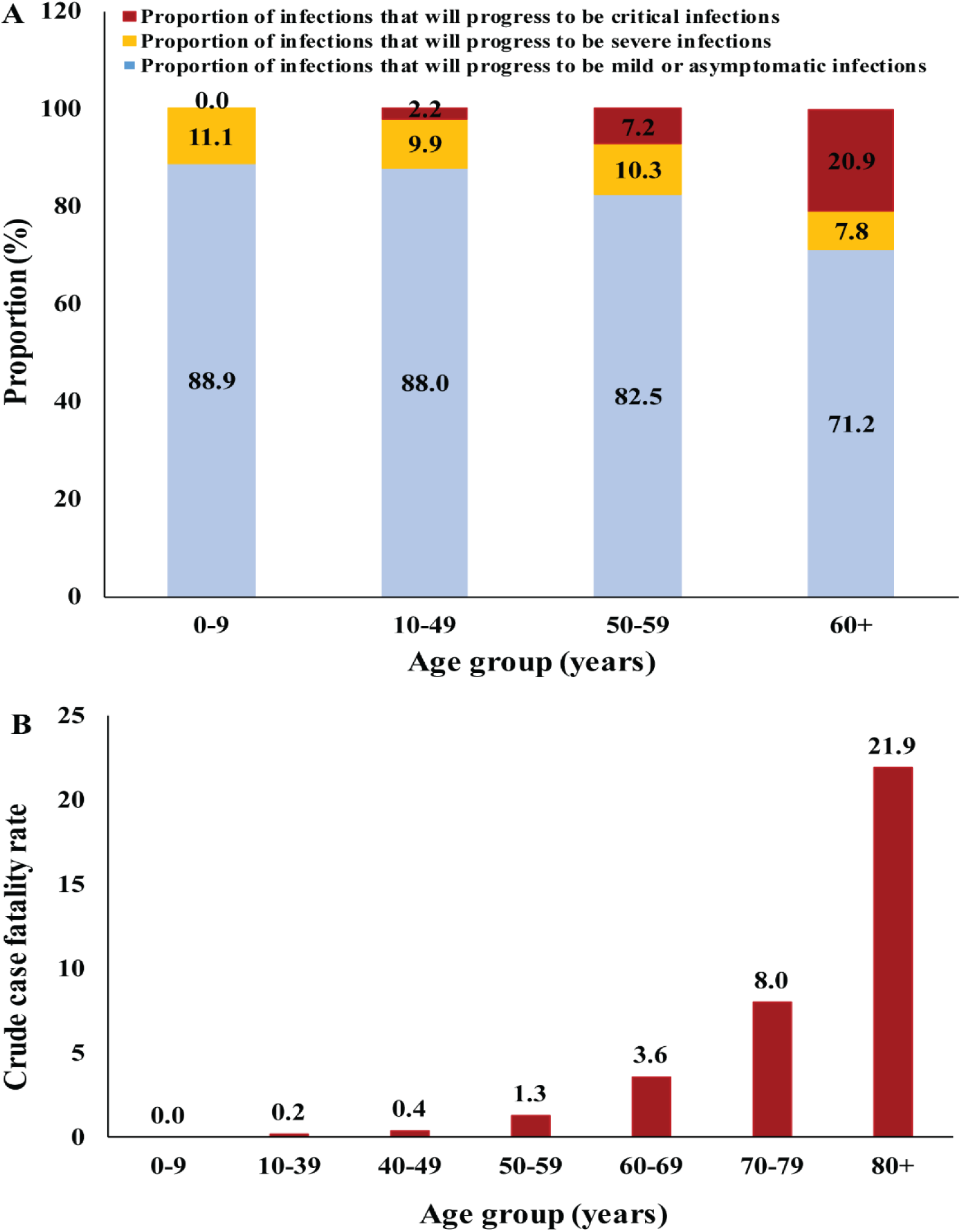
Key attributes of SARS-CoV-2 disease progression stratified by age. Distribution by age of: A) the proportion of infections that will progress to be mild or asymptomatic, severe, or critical [2-4] and B) crude case fatality rate [5, 6].

## 1. Mathematical model structure

We applied a recently developed deterministic compartmental mathematical model that describes the severe acute respiratory syndrome coronavirus 2 (SARS-CoV-2) transmission dynamics and disease progression in a population [1], to all countries and territories with a population of at least one million, as of 2020 [7]. An illustration of the basic model structure can be found in Figure S3. The model stratifies the population into compartments based on age (0-9, 10-19, 20- 29,…, ≥80 years), infection status (uninfected, infected), infection stage (mild, severe, critical), and disease stage (severe, critical).

**Figure S3:**
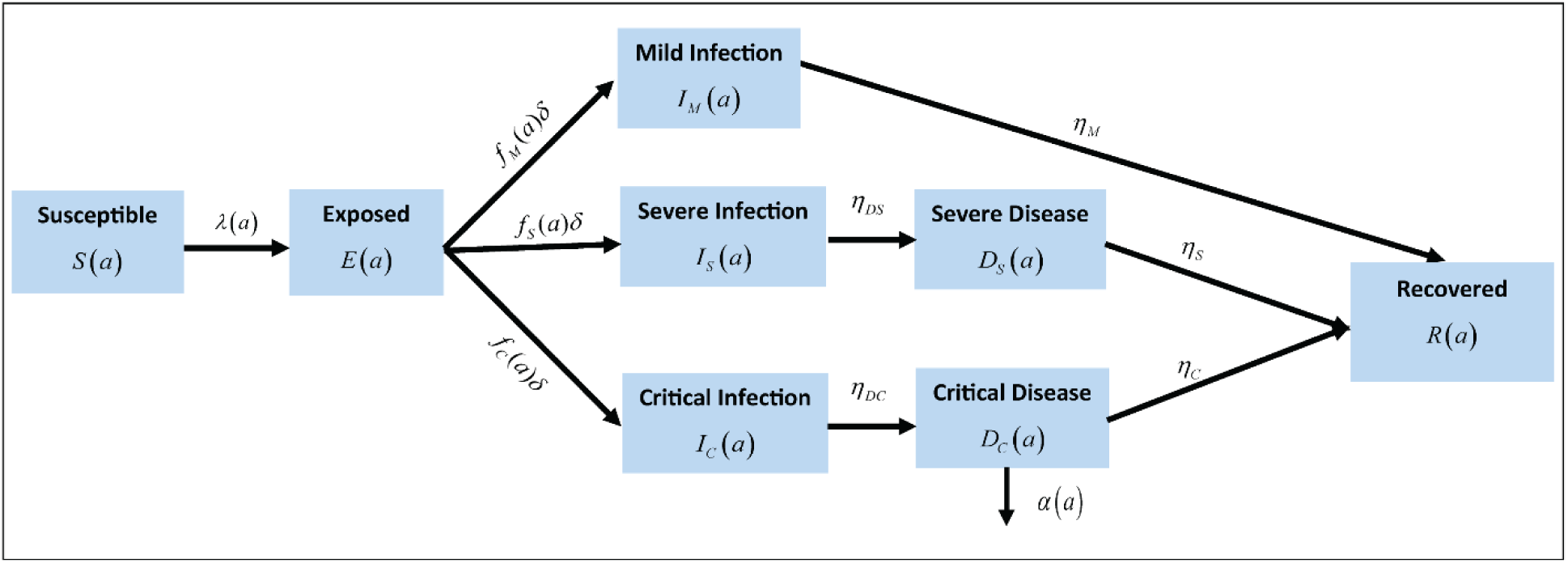
**Schematic diagram illustrating the basic structure of the SARS-CoV-2 model.**

A system of coupled nonlinear differential equations is used to describe SARS-CoV-2 transmission dynamics. Nine age cohorts were considered (*a* = 1, 2,…, 9), each representing a ten-year age band apart from the last cohort which includes individuals aged ≥80 years. The age-specific distribution for each country/territory, as of the year 2020, was obtained from the United Nations World Population Prospects database [7]. All disease-related mortality was assumed to occur in individuals that are in the critical disease stage, as informed by the China outbreak data [5].

The epidemic dynamics in the first age group was described using:

### Population aged 0-9 years

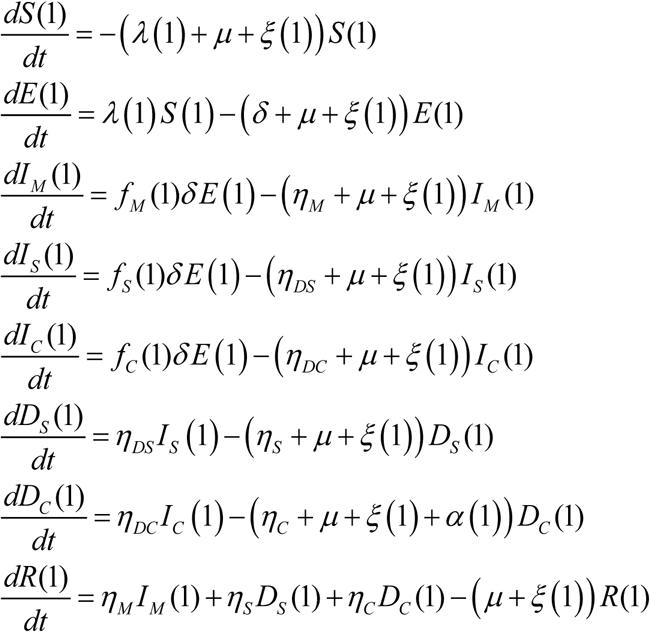

and in subsequent age groups, using:

### Populations aged 10+ years

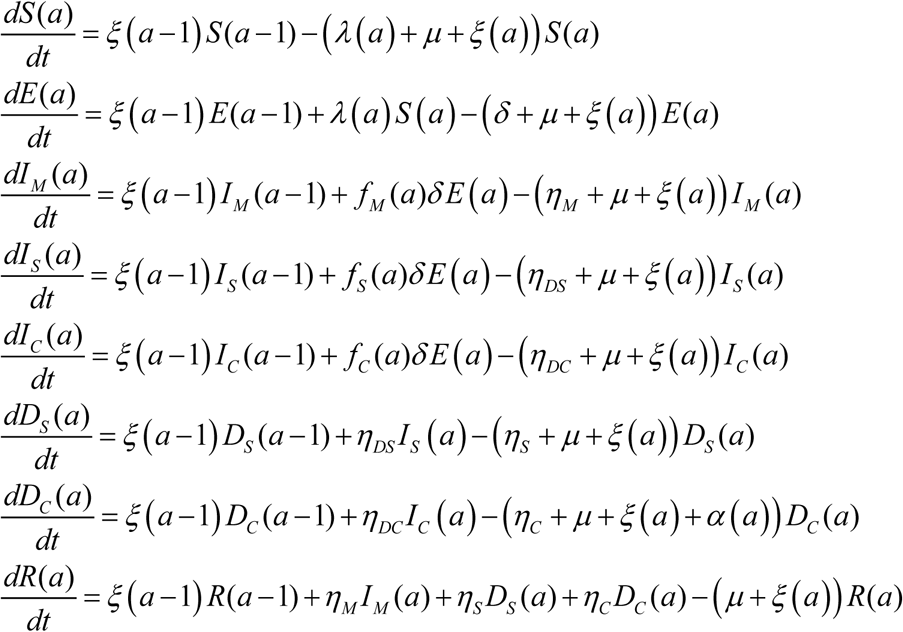

The definitions of the population variables and symbols used in the equations are listed in Table S1.

**Table S1:**
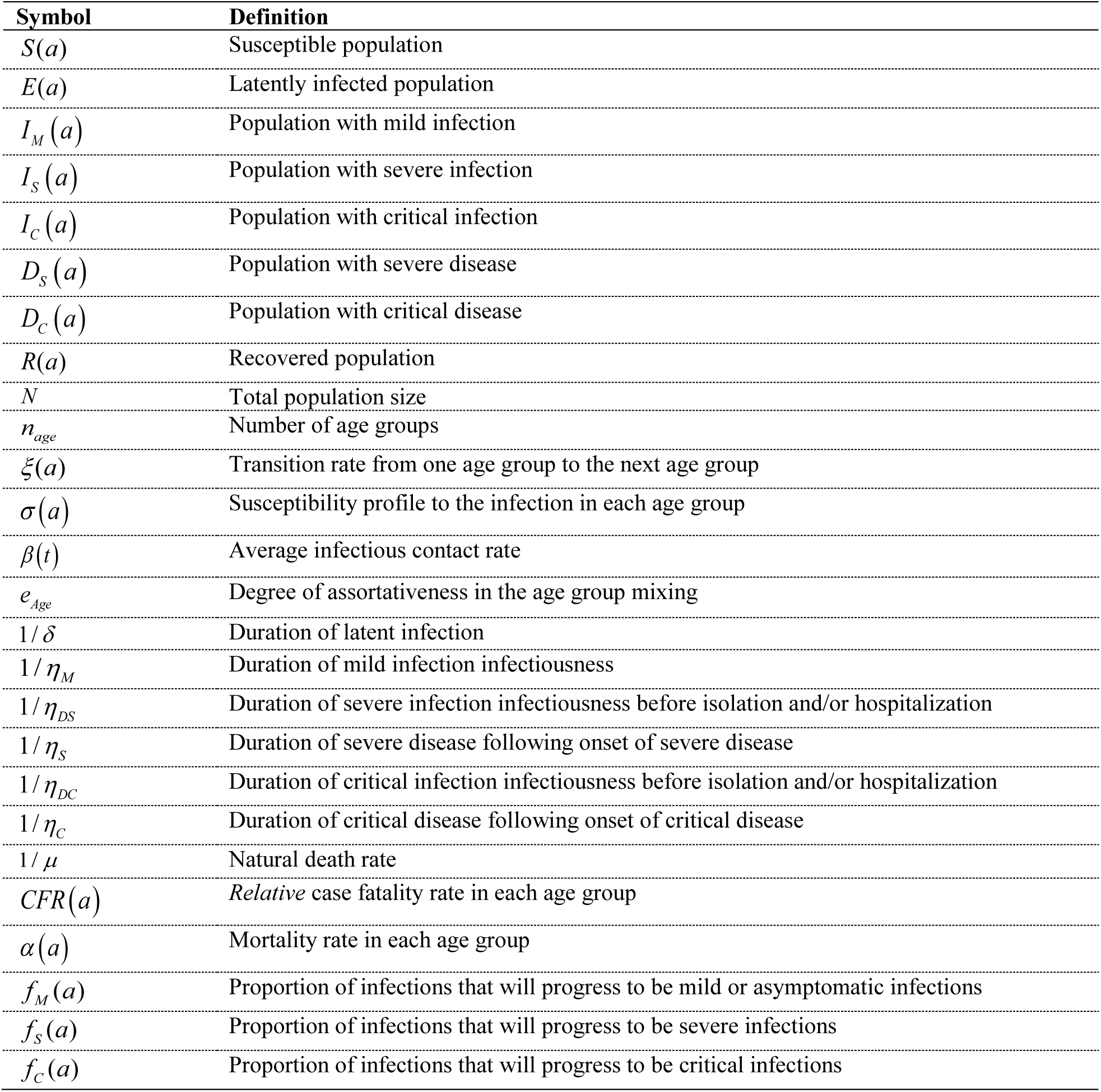
Definitions of population variables and symbols used in the model.

| Symbol | Definition |
| --- | --- |
| $S(a)$ | Susceptible population |
| $E(a)$ | Latently infected population |
| $I_M(a)$ | Population with mild infection |
| $I_S(a)$ | Population with severe infection |
| $I_C(a)$ | Population with critical infection |
| $D_S(a)$ | Population with severe disease |
| $D_C(a)$ | Population with critical disease |
| $R(a)$ | Recovered population |
| $N$ | Total population size |
| $n_{age}$ | Number of age groups |
| $\xi(a)$ | Transition rate from one age group to the next age group |
| $\sigma(a)$ | Susceptibility profile to the infection in each age group |
| $\beta(t)$ | Average infectious contact rate |
| $e_{Age}$ | Degree of assortativeness in the age group mixing |
| $1/\delta$ | Duration of latent infection |
| $1/\eta_M$ | Duration of mild infection infectiousness |
| $1/\eta_{DS}$ | Duration of severe infection infectiousness before isolation and/or hospitalization |
| $1/\eta_S$ | Duration of severe disease following onset of severe disease |
| $1/\eta_{DC}$ | Duration of critical infection infectiousness before isolation and/or hospitalization |
| $1/\eta_C$ | Duration of critical disease following onset of critical disease |
| $1/\mu$ | Natural death rate |
| $CFR(a)$ | Relative case fatality rate in each age group |
| $\alpha(a)$ | Mortality rate in each age group |
| $f_M(a)$ | Proportion of infections that will progress to be mild or asymptomatic infections |
| $f_S(a)$ | Proportion of infections that will progress to be severe infections |
| $f_C(a)$ | Proportion of infections that will progress to be critical infections |

The force of infection (hazard rate) for each susceptible population in each age group *S*(*a*) is given by

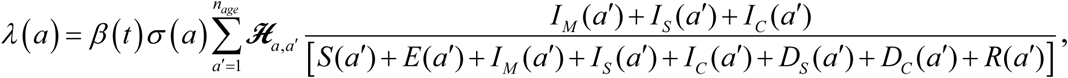

where *β* (*t*) is the overall infectious contact rate per day, *σ* (*a*) is the susceptibility profile to the infection in each age group, and ℋ_*a,a*′_ is the mixing matrix which provides the probability that an individual in the *a* age group will mix with (that is contact) an individual in the *a* ′ age group. The mixing matrix is given by

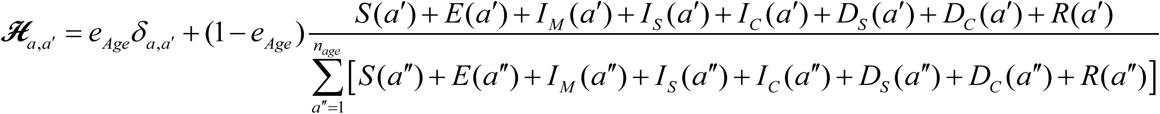

Here, *δ*_*a,a*′_ is the identity matrix, and *e*_*Age*_ ∈ [0,1] is the degree of assortativeness in the mixing. At the extreme *e*_*Age*_ = 0, the mixing is fully proportional, while at the other extreme *e*_*Age*_ = 1, the mixing is fully assortative, that is individuals mix only with members in their own age group.

## 2. Parameter values

The input parameters of the model were chosen based on current empirical data for SARS-CoV- 2 natural history and epidemiology. The parameter values are listed in Table S2.

**Table S2:** Model assumptions in terms of parameter values.

| Parameter | Symbol | Value | Justification |
| --- | --- | --- | --- |
| Duration of latent infection | $1/\delta$ | 3.69 days | Based on existing estimate [8] and based on a median incubation period of 5.1 days [9] adjusted by observed viral load among infected persons [10] and reported transmission before onset of symptoms [11] |
| Duration of infectiousness | $1/\eta_M$ ;<br>$1/\eta_{DS}$ ;<br>$1/\eta_{DC}$ | 3.48 days | Based on existing estimate [8] and based on observed time to recovery among persons with mild infection [4, 8] and observed viral load in infected persons [10, 11] |
| Duration of severe disease following onset of severe disease | $1/\eta_S$ | 28 days | Observed duration from onset of severe disease to recovery [4] |
| Duration of hospitalization for critical infection | $1/\eta_C$ | 28 days | Observed duration from onset of critical disease to recovery [4] |
| Life expectancy for each country | $1/\mu$ | Table S3 | United Nations World Population Prospects database [7] |
| Relative case fatality rate in each age group | $CFR(a)$ | | Observed crude case fatality rate based on China data [5, 6] |
| Age 0-9 years |  | 0% |  |
| Age 10-39 years |  | 0.2% |  |
| Age 40-49 years |  | 0.4% |  |
| Age 50-59 years |  | 1.3% |  |
| Age 60-69 years |  | 3.6% |  |
| Age 70-79 years |  | 8.0% |  |
| Age 80+ years |  | 21.9% |  |
| Proportion of infections that will progress to be mild or asymptomatic infections | $f_M(a)$ | | Observed proportion of infections that eventually develop mild or asymptomatic in China [2-4] |
| Age 0-9 years |  | 88.9% |  |
| Age 10-49 years |  | 88.0% |  |
| Age 50-59 years |  | 82.5% |  |
| Age 60+ years |  | 71.2% |  |
| Proportion of infections that will progress to be severe infections | $f_S(a)$ | | Observed proportion of infections that eventually develop severe disease in China [2-4] |
| Age 0-9 years |  | 11.1% |  |
| Age 10-49 years |  | 9.9% |  |
| Age 50-59 years |  | 10.3% |  |
| Age 60+ years |  | 7.8% |  |
| Proportion of infections that will progress to be critical infections | $f_C(a)$ | | Observed proportion of infections that eventually develop critical disease in China [2-4] |
| Age 0-9 years |  | 0.0% |  |
| Age 10-49 years |  | 2.2% |  |
| Age 50-59 years |  | 7.2% |  |
| Age 60+ years |  | 20.9% |  |
| Susceptibility profile to the infection in each age group | $\sigma(a)$ | | Estimates based on fitting the epidemic in China [1] |
| Age 0-9 years |  | 0.05 |  |
| Age 10-19 years |  | 0.05 |  |
| Age 20-29 years |  | 0.32 |  |
| Age 30-39 years |  | 0.53 |  |
| Age 40-49 years |  | 0.65 |  |
| Age 50-59 years |  | 0.74 |  |
| Age 60-69 years |  | 0.93 |  |
| Age 70-79 years |  | 0.88 |  |
| Age 80+ years |  | 0.83 |  |
| Overall infectious contact rate | $\beta(t)$ | 0.59 contacts per day | Estimate based on fitting the epidemic in China [1] |
| Degree of assortativeness in the age group mixing | $e_{Age}$ | 0.004 | Estimate based on fitting the epidemic in China [1] |

## 3. The basic reproduction number *R*_*0*_

Using the second generation matrix method described by Heffernan *et al*. [12], the basic reproduction number was derived to be: 

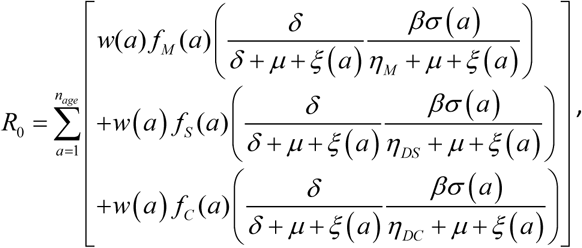

where *w*(*a*) is the proportion of the population in each age group.

**Table S3:** **Model estimates for key SARS-CoV-2 epidemiologic indicators for countries and territories with a population of at least one million [7] in the World Health Organization African Region (AFRO)**.

| African Region | Total population [7] | Median age [7] | Life expectancy [7] | $R_0^a$ | Infections per 100 persons<sup>a</sup> | Deaths per 100 persons<sup>a</sup> | Mild infections per 100 persons<sup>a</sup> | Severe and/or critical disease cases per 100 persons<sup>a</sup> | Day at peak incidence<sup>a</sup> |
| --- | --- | --- | --- | --- | --- | --- | --- | --- | --- |
| Country | Thousands | Years | Years | N (95% UI) | N (95% UI) | N (95% UI) | N (95% UI) | N (95% UI) | N (95% UI) |
| Algeria | 43,851,043 | 28.5 | 77.5 | 1.45 (1.35-2.21) | 45.0 (35.2-52.6) | 1.4 (1.2-1.6) | 37.0 (29.5-44.3) | 7.4 (5.7-8.3) | 217.5 (206.0-246.0) |
| Angola | 32,866,268 | 16.7 | 62.2 | 0.98 (0.88-1.46) | 1.5 (0.0-23.4) | 0.0 (0.0-0.5) | 1.0 (0.0-20.0) | 0.2 (0.0-3.4) | 697.5 (518.0-700<sup>b</sup>) |
| Benin | 12,123,198 | 18.8 | 62.8 | 1.05 (0.98-1.61) | 19.5 (0.1-29.8) | 0.6 (0.0-0.7) | 17.0 (0.1-25.4) | 3.3 (0.0-4.5) | 675.0 (360.0-700<sup>b</sup>) |
| Botswana | 2,351,625 | 24.0 | 69.9 | 1.25 (1.18-1.94) | 35.0 (23.6-43.6) | 0.9 (0.7-1.1) | 29.0 (19.9-37.0) | 5.3 (3.7-6.6) | 235.0 (212.0-306.0) |
| Burkina Faso | 20,903,278 | 17.6 | 63.0 | 0.98 (0.91-1.51) | 2.5 (0.0-25.6) | 0.1 (0.0-0.5) | 1.5 (0.0-21.9) | 0.3 (0.0-3.8) | 700<sup>b</sup> (453.0-700<sup>b</sup>) |
| Burundi | 11,890,781 | 17.3 | 62.7 | 1.03 (0.90-1.49) | 2.5 (0.0-24.8) | 0.1 (0.0-0.5) | 1.5 (0.0-21.1) | 0.3 (0.0-3.6) | 700<sup>b</sup> (454.0-700<sup>b</sup>) |
| Cameroon | 26,545,864 | 18.7 | 60.3 | 1.05 (0.96-1.59) | 2.5 (0.0-29.2) | 0.1 (0.0-0.6) | 17.5 (0.0-24.9) | 0.3 (0.0-4.3) | 675.0 (397.0-700<sup>b</sup>) |
| Cent. Afr. Rep. | 4,829,764 | 17.6 | 54.4 | 0.98 (0.91-1.50) | 2.5 (0.0-24.7) | 0.1 (0.0-0.6) | 1.5 (0.0-21.0) | 0.3 (0.0-3.7) | 700<sup>b</sup> (420.0-700<sup>b</sup>) |
| Chad | 16,425,859 | 16.6 | 55.2 | 0.92 (0.86-1.43) | 1.0 (0.0-22.0) | 0.0 (0.0-0.5) | 1.0 (0.0-18.7) | 0.2 (0.0-3.2) | 697.5 (532.0-700<sup>b</sup>) |
| Congo | 5,518,092 | 19.2 | 65.2 | 1.15 (1.01-1.66) | 23.0 (2.5-31.7) | 0.5 (0.1-0.7) | 19.0 (2.2-27.0) | 3.4 (0.4-4.7) | 405.0 (316.0-700<sup>b</sup>) |
| Côte d'Ivoire | 26,378,275 | 18.9 | 58.8 | 1.05 (0.97-1.60) | 22.5 (0.0-29.6) | 0.6 (0.0-0.7) | 1.5 (0.0-25.2) | 3.3 (0.0-4.4) | 675.0 (389.0-700<sup>b</sup>) |
| D. Rep. Congo | 89,561,404 | 17.0 | 61.6 | 0.98 (0.91-1.51) | 2.5 (0.0-25.4) | 0.1 (0.0-0.6) | 1.5 (0.0-21.6) | 0.3 (0.0-3.8) | 690.0 (497.0-700<sup>b</sup>) |
| Equ. Guinea | 1,402,985 | 22.3 | 59.8 | 1.15 (1.05-1.74) | 27.0 (14.7-36.7) | 0.5 (0.3-0.6) | 23.0 (12.6-31.6) | 3.8 (2.1-5.2) | 310.0 (259.0-534.0) |
| Eritrea | 3,546,427 | 19.2 | 67.5 | 1.15 (1.01-1.67) | 23.0 (6.1-31.7) | 0.7 (0.2-0.9) | 19.0 (5.2-26.9) | 3.5 (1.0-4.9) | 345.0 (303.0-700<sup>b</sup>) |
| Ethiopia | 114,963,583 | 19.5 | 67.8 | 1.05 (0.99-1.63) | 22.5 (0.0-31.5) | 0.7 (0.0-0.8) | 19.5 (0.0-26.8) | 3.8 (0.0-4.7) | 675.0 (402.0-700<sup>b</sup>) |
| Eswatini | 1,160,164 | 20.7 | 61.1 | 1.15 (1.04-1.72) | 25.0 (13.7-34.5) | 0.7 (0.4-0.8) | 23.0 (11.6-29.4) | 3.8 (2.1-5.1) | 330.0 (261.0-547.0) |
| Gabon | 2,225,728 | 22.5 | 67.0 | 1.25 (1.11-1.83) | 29.0 (18.4-39.1) | 0.7 (0.5-0.9) | 25.0 (15.6-33.3) | 4.4 (2.8-5.8) | 275.0 (239.0-404.0) |
| Gambia | 2,416,664 | 17.8 | 63.3 | 1.03 (0.92-1.52) | 2.5 (0.0-25.5) | 0.1 (0.0-0.5) | 1.5 (0.0-21.8) | 0.3 (0.0-3.7) | 675.0 (383.0-700<sup>b</sup>) |
| Ghana | 31,072,945 | 21.5 | 64.9 | 1.15 (1.09-1.79) | 29.0 (17.7-37.7) | 0.7 (0.5-0.8) | 25.0 (15.0-32.1) | 4.4 (2.7-5.6) | 350.0 (298.0-538.0) |
| Guinea | 13,132,792 | 18.0 | 62.6 | 1.03 (0.93-1.53) | 2.5 (0.0-26.4) | 0.1 (0.0-0.6) | 2.5 (0.0-22.4) | 0.3 (0.0-3.9) | 700<sup>b</sup> (423.0-700<sup>b</sup>) |
| Guinea-Bissau | 1,967,998 | 18.8 | 59.4 | 1.05 (0.97-1.60) | 19.0 (0.1-29.0) | 0.5 (0.0-0.6) | 17.0 (0.1-24.7) | 2.9 (0.0-4.3) | 675.0 (326.0-700<sup>b</sup>) |
| Kenya | 53,771,300 | 20.1 | 67.5 | 1.15 (1.01-1.67) | 25.0 (0.6-33.5) | 0.5 (0.0-0.6) | 21.0 (0.5-28.6) | 3.4 (0.1-4.9) | 425.0 (361.0-700<sup>b</sup>) |
| Lesotho | 2,142,252 | 24.0 | 55.7 | 1.25 (1.18-1.95) | 37.0 (23.9-43.9) | 1.0 (0.7-1.2) | 29.0 (20.2-37.2) | 5.6 (3.8-6.7) | 235.0 (210.0-299.0) |
| Liberia | 5,057,677 | 19.4 | 65.0 | 1.15 (1.01-1.67) | 23.0 (4.0-31.9) | 0.6 (0.1-0.7) | 21.0 (3.4-27.1) | 3.4 (0.6-4.8) | 375.0 (312.0-700<sup>b</sup>) |
| Madagascar | 27,691,019 | 19.6 | 68.2 | 1.05 (1.00-1.66) | 23.0 (0.6-32.1) | 0.6 (0.0-0.7) | 19.0 (0.5-27.3) | 3.4 (0.1-4.8) | 425.0 (355.0-700<sup>b</sup>) |
| Malawi | 19,129,955 | 18.1 | 65.6 | 1.03 (0.92-1.53) | 2.5 (0.0-26.7) | 0.1 (0.0-0.6) | 2.5 (0.0-22.8) | 0.3 (0.0-3.9) | 700<sup>b</sup> (434.0-700<sup>b</sup>) |
| Mali | 20,250,834 | 16.3 | 60.5 | 0.98 (0.87-1.44) | 1.0 (0.0-22.3) | 0.0 (0.0-0.5) | 1.0 (0.0-19.0) | 0.2 (0.0-3.3) | 697.5 (526.0-700<sup>b</sup>) |
| Mauritania | 4,649,660 | 20.1 | 65.6 | 1.15 (1.03-1.70) | 25.0 (11.1-33.6) | 0.6 (0.3-0.8) | 21.0 (9.4-28.7) | 3.8 (1.7-5.0) | 375.0 (294.0-642.0) |
| Mauritius | 1,271,767 | 37.5 | 75.5 | 1.95 (1.72-2.80) | 63.5 (56.7-69.9) | 2.8 (2.6-3.1) | 52.5 (46.6-57.7) | 11.3 (10.1-12.2) | 116.0 (115.0-117.0) |
| Mozambique | 31,255,435 | 17.6 | 62.1 | 1.03 (0.92-1.51) | 2.5 (0.0-25.8) | 0.1 (0.0-0.6) | 1.5 (0.0-22.0) | 0.3 (0.0-3.8) | 700<sup>b</sup> (462.0-700<sup>b</sup>) |
| Namibia | 2,540,916 | 21.8 | 64.9 | 1.15 (1.08-1.79) | 31.0 (17.0-37.6) | 0.7 (0.5-0.9) | 19.0 (14.4-32) | 4.4 (2.6-5.6) | 290.0 (253.0-457.0) |
| Niger | 24,206,636 | 15.2 | 63.6 | 0.92 (0.83-1.37) | 0.3 (0.0-17.0) | 0.0 (0.0-0.4) | 0.3 (0.0-14.4) | 0.1 (0.0-2.5) | 697.5 (624.0-700<sup>b</sup>) |
| Nigeria | 206,139,587 | 18.1 | 55.8 | 1.03 (0.95-1.57) | 2.5 (0.0-28.6) | 0.1 (0.0-0.6) | 2.5 (0.0-24.4) | 0.5 (0.0-4.3) | 705.0 (459.0-700<sup>b</sup>) |
| Rwanda | 12,952,209 | 20.0 | 70.0 | 1.15 (1.03-1.69) | 25.0 (6.5-33.5) | 0.6 (0.2-0.8) | 19.0 (5.5-28.5) | 3.7 (1.0-5.0) | 405.0 (320.0-700<sup>b</sup>) |
| Senegal | 16,743,930 | 18.5 | 68.9 | 1.05 (0.96-1.59) | 22.5 (0.0-29.0) | 0.6 (0.0-0.7) | 1.5 (0.0-24.7) | 3.3 (0.0-4.3) | 675.0 (382.0-700<sup>b</sup>) |
| Sierra Leone | 7,976,985 | 19.4 | 55.9 | 1.05 (0.99-1.64) | 21.0 (0.4-31.1) | 0.5 (0.0-0.7) | 19.0 (0.4-26.5) | 3.2 (0.1-4.6) | 425.0 (338.0-700<sup>b</sup>) |
| South Africa | 59,308,690 | 27.6 | 64.9 | 1.45 (1.32-2.16) | 45.0 (33.7-52.0) | 1.2 (1.0-1.4) | 37.0 (28.4-44.0) | 7.1 (5.4-8.0) | 232.5 (217.0-268.0) |

| **African Region** | **Total population [7]** | **Median age [7]** | **Life expectancy [7]** | **$R_0^a$** | **Infections per 100 persons<sup>a</sup>** | **Deaths per 100 persons<sup>a</sup>** | **Mild infections per 100 persons<sup>a</sup>** | **Severe and/or critical disease cases per 100 persons<sup>a</sup>** | **Day at peak incidence<sup>a</sup>** |
| --- | --- | --- | --- | --- | --- | --- | --- | --- | --- |
| **Country** | **Thousands** | **Years** | **Years** | **N (95% UI)** | **N (95% UI)** | **N (95% UI)** | **N (95% UI)** | **N (95% UI)** | **N (95% UI)** |
| South Sudan | 11,193,729 | 19.0 | 58.7 | 1.05 (0.98-1.62) | 21.0 (0.2-30.3) | 0.6 (0.0-0.7) | 17.0 (0.1-25.8) | 3.2 (0.0-4.5) | 675.0 (353.0-700<sup>b</sup>) |
| Togo | 8,278,737 | 19.4 | 62.1 | 1.05 (1.00-1.65) | 23.0 (1.1-31.6) | 0.6 (0.0-0.7) | 19.0 (1.0-27.0) | 3.4 (0.2-4.7) | 375.0 (330.0-700<sup>b</sup>) |
| Uganda | 45,741,000 | 16.7 | 64.4 | 0.92 (0.86-1.42) | 0.5 (0.0-22.0) | 0.0 (0.0-0.4) | 0.5 (0.0-18.8) | 0.1 (0.0-3.2) | 697.5 (575.0-700<sup>b</sup>) |
| Tanzania | 59,734,213 | 18.0 | 66.4 | 0.98 (0.94-1.55) | 2.5 (0.0-27.7) | 0.1 (0.0-0.6) | 2.5 (0.0-23.6) | 0.5 (0.0-4.1) | 705.0 (443.0-700<sup>b</sup>) |
| Zambia | 18,383,956 | 17.6 | 64.7 | 1.03 (0.89-1.48) | 2.5 (0.0-24.9) | 0.1 (0.0-0.5) | 1.5 (0.0-21.3) | 0.3 (0.0-3.6) | 690.0 (472.0-700<sup>b</sup>) |
| Zimbabwe | 14,862,927 | 18.7 | 62.2 | 1.05 (0.97-1.60) | 22.5 (0.0-29.3) | 0.6 (0.0-0.7) | 16.5 (0.0-25.0) | 3.3 (0.0-4.3) | 675.0 (377.0-700<sup>b</sup>) |
| **AFRO** | **1,118,418,151** | **18.9** | **63.3<sup>c</sup>** | **1.05 (0.93-1.95)<sup>c</sup>** | **22.5 (0.25-63.5)<sup>c</sup>** | **0.5 (0.0-2.8)<sup>c</sup>** | **17.0 (0.25-52.5)<sup>c</sup>** | **3.3 (0.0-11.3)<sup>c</sup>** | **675 (116.0-700<sup>b</sup>)<sup>c</sup>** |

Cent Afr Rep, Central African Republic. D Rep Congo, Democratic Republic of the Congo. Equ Guinea, Equatorial Guinea. N, number. NA, not applicable. Tanzania, United Republic of Tanzania. UI, uncertainty interval.
<sup>a</sup>For each epidemiologic indicator, the table reports the most probable estimate and the 95% uncertainty interval. For a number of countries, $R_0$ was close to 1 in some of the uncertainty runs leading to an estimated time for peak incidence beyond the simulation duration of two years (delayed epidemic peak). For these countries, the upper limit of the uncertainty interval was set at 700 days, that is the end day of the simulation. Uncertainty runs in which $R_0$ was less than 1 led to no epidemic emergence, and thus were excluded from further analysis. Reported ranges for the epidemic indicators did not include these runs, except for the range of $R_0$ .
<sup>b</sup>The estimated time for peak incidence was beyond the simulation duration of two years. The upper limit of the uncertainty interval was set at 700 days, that is the end day of the simulation.
<sup>c</sup>Estimates are for the median (and range).

**Table S4:** **Model estimates for key SARS-CoV-2 epidemiologic indicators for countries and territories with a population of at least one million [7] in the World Health Organization Region of the Americas (AMRO)**.

| Region of the Americas | Total population [7] | Median age [7] | Life expectancy [7] | $R_0^a$ | Infections per 100 persons<sup>a</sup> | Deaths per 100 persons<sup>a</sup> | Mild infections per 100 persons<sup>a</sup> | Severe and/or critical disease cases per 100 persons<sup>a</sup> | Day at peak incidence<sup>a</sup> |
| --- | --- | --- | --- | --- | --- | --- | --- | --- | --- |
| Country | Thousands | Years | Years | N (95% UI) | N (95% UI) | N (95% UI) | N (95% UI) | N (95% UI) | N (95% UI) |
| Argentina | 45,195,777 | 31.5 | 77.2 | 1.65 (1.51-2.47) | 53.0 (44.8-60.0) | 2.3 (2.0-2.5) | 45.5 (37.0-49.8) | 9.1 (7.8-10.2) | 180.5 (174.0-188.0) |
| Bolivia | 11,673,029 | 25.6 | 72.4 | 1.45 (1.28-2.10) | 41.0 (30.3-48.6) | 1.5 (1.2-1.6) | 33.0 (25.3-40.8) | 6.8 (5.0-7.8) | 227.5 (207.0-262.0) |
| Brazil | 212,559,409 | 33.5 | 76.6 | 1.75 (1.57-2.57) | 57.0 (49.3-64.3) | 2.1 (2.0-2.4) | 49.5 (41.0-53.7) | 9.7 (8.4-10.6) | 183.5 (180.0-190.0) |
| Canada | 37,742,157 | 41.1 | 83.0 | 2.05 (1.86-3.02) | 69.5 (63.0-74.5) | 3.7 (3.6-4.1) | 56.5 (51.1-60.7) | 13.1 (11.9-13.8) | 131.3 (127.0-133.0) |
| Chile | 19,116,209 | 35.3 | 80.7 | 1.85 (1.66-2.71) | 61.5 (54.0-67.7) | 2.6 (2.4-2.9) | 51.5 (44.5-56.1) | 10.7 (9.5-11.6) | 146.9 (145.0-148.0) |
| Colombia | 50,882,884 | 31.3 | 77.9 | 1.65 (1.50-2.46) | 55.0 (45.4-61.1) | 2.0 (1.8-2.2) | 45.5 (37.7-51.1) | 9.1 (7.7-10.0) | 182.5 (176.0-191.0) |
| Costa Rica | 5,094,114 | 33.5 | 80.9 | 1.75 (1.58-2.59) | 59.0 (49.9-64.6) | 2.3 (2.1-2.5) | 48.5 (41.4-53.8) | 9.9 (8.6-10.8) | 144.3 (142.0-148.0) |
| Cuba | 11,326,616 | 42.2 | 79.2 | 2.05 (1.84-2.99) | 68.5 (62.4-74.1) | 3.3 (3.2-3.7) | 56.5 (50.9-60.7) | 12.5 (11.5-13.4) | 122.3 (120.0-125.0) |
| Dom. Rep. | 10,847,904 | 28.0 | 74.7 | 1.45 (1.37-2.24) | 47.0 (36.5-53.8) | 1.6 (1.4-1.8) | 39.0 (30.4-45.1) | 7.7 (6.1-8.7) | 196.5 (184.0-215.0) |
| Ecuador | 17,643,060 | 27.9 | 77.7 | 1.45 (1.36-2.24) | 47.0 (36.4-53.7) | 1.7 (1.4-1.8) | 39.0 (30.4-45.1) | 7.7 (6.0-8.6) | 202.5 (191.0-224.0) |
| El Salvador | 6,486,201 | 27.6 | 74.1 | 1.45 (1.37-2.25) | 47.0 (36.8-54.0) | 1.7 (1.5-1.9) | 39.0 (30.6-45.2) | 7.7 (6.2-8.8) | 187.5 (178.0-207.0) |
| Guatemala | 17,915,567 | 22.9 | 75.1 | 1.25 (1.14-1.88) | 35.0 (21.3-41.3) | 1.0 (0.7-1.1) | 29.0 (18.0-35.1) | 5.0 (3.4-6.3) | 305.0 (262.0-410.0) |
| Haiti | 11,402,533 | 24.0 | 65.0 | 1.35 (1.19-1.96) | 35.0 (24.7-44.3) | 1.1 (0.8-1.2) | 31.0 (20.8-37.5) | 5.6 (3.9-6.8) | 265.0 (233.0-332.0) |
| Honduras | 9,904,608 | 24.3 | 75.9 | 1.35 (1.20-1.98) | 37.0 (25.8-45.6) | 1.0 (0.8-1.2) | 31.0 (21.7-38.7) | 5.9 (4.1-6.9) | 255.0 (227.0-318.0) |
| Jamaica | 2,961,161 | 30.7 | 74.9 | 1.65 (1.48-2.43) | 53.0 (43.9-59.9) | 2.0 (1.8-2.2) | 45.5 (36.5-50.0) | 9.1 (7.5-9.9) | 155.5 (150.0-162.0) |
| Mexico | 128,932,753 | 29.2 | 75.4 | 1.55 (1.41-2.31) | 49.0 (39.5-56.2) | 1.7 (1.5-1.9) | 40.5 (32.9-47.2) | 8.0 (6.5-9.1) | 214.5 (204.0-234.0) |
| Nicaragua | 6,624,554 | 26.5 | 75.2 | 1.45 (1.28-2.11) | 41.0 (31.1-49.8) | 1.2 (1.0-1.4) | 35.0 (26.1-42.1) | 6.5 (5.0-7.7) | 212.5 (198.0-250.0) |
| Panama | 4,314,768 | 29.7 | 79.1 | 1.55 (1.42-2.34) | 49.0 (39.9-56.4) | 1.8 (1.6-2.0) | 40.5 (33.2-47.2) | 8.3 (6.7-9.2) | 171.0 (163.0-182.0) |
| Paraguay | 7,132,530 | 26.3 | 74.6 | 1.45 (1.29-2.13) | 43.0 (31.8-50.3) | 1.4 (1.1-1.6) | 35.0 (26.6-42.3) | 7.1 (5.2-8) | 212.5 (196.0-244.0) |
| Peru | 32,971,846 | 31.0 | 77.4 | 1.65 (1.47-2.41) | 53.0 (43.3-59.4) | 2.0 (1.7-2.1) | 43.5 (36.1-49.7) | 8.7 (7.3-9.7) | 185.0 (177.0-195.0) |
| Puerto Rico | 2,860,840 | 44.5 | 80.7 | 2.28 (1.99-2.61) | 70.5 (64.0-74.7) | 4.2 (4.0-4.6) | 56.5 (51.5-60.4) | 13.5 (12.5-14.3) | 106.3 (103.0-110.0) |
| Trin. and Tob. | 1,399,491 | 36.2 | 73.9 | 1.85 (1.66-2.72) | 61.5 (54.0-67.6) | 2.5 (2.3-2.8) | 51.5 (44.5-56.1) | 10.7 (9.4-11.6) | 121.9 (121.0-123.0) |
| USA | 331,002,647 | 38.3 | 79.1 | 1.95 (1.76-2.86) | 64.5 (57.9-70.2) | 3.4 (3.2-3.8) | 52.5 (47.1-57.4) | 12.1 (10.9-12.9) | 158.9 (156.0-160.0) |
| Uruguay | 3,473,727 | 35.8 | 78.4 | 1.85 (1.66-2.71) | 60.5 (53.3-66.6) | 3.2 (2.8-3.3) | 49.5 (43.5-54.8) | 10.9 (9.7-11.9) | 130.9 (129.0-132.0) |
| Venezuela | 28,435,943 | 29.6 | 72.3 | 1.55 (1.41-2.32) | 49.0 (39.2-55.5) | 1.8 (1.5-2.0) | 40.5 (32.6-46.4) | 8.0 (6.6-9.1) | 195.0 (185.0-210.0) |
| **AMRO** | **1,017,900,328** | **30.7** | **76.6<sup>c</sup>** | **1.65 (1.25-2.28)<sup>c</sup>** | **53.0 (35.0-70.5)<sup>c</sup>** | **2.0 (1.0-4.2)<sup>c</sup>** | **43.5 (29.0-56.5)<sup>c</sup>** | **8.7 (5.0-13.5)<sup>c</sup>** | **183.5 (106.3-305.0)<sup>c</sup>** |

Dom Rep, Dominican Republic. N, number. Trin. and Tob., Trinidad and Tobago. UI, uncertainty interval. USA, United States of America.
<sup>a</sup>For each epidemiologic indicator, the table reports the most probable estimate and the 95% uncertainty interval. For a number of countries, $R_0$ was close to 1 in some of the uncertainty runs leading to an estimated time for peak incidence beyond the simulation duration of two years (delayed epidemic peak). For these countries, the upper limit of the uncertainty interval was set at 700 days, that is the end day of the simulation. Uncertainty runs in which $R_0$ was less than 1 led to no epidemic emergence, and thus were excluded from further analysis. Reported ranges for the epidemic indicators did not include these runs, except for the range of $R_0$ .
<sup>c</sup>Estimates are for the median (and range).

**Table S5:** **Model estimates for key SARS-CoV-2 epidemiologic indicators for countries and territories with a population of at least one million [7] in the World Health Organization Eastern Mediterranean Region (EMRO)**.

| Eastern Mediterranean Region | Total population [7] | Median age [7] | Life expectancy [7] | $R_0^a$ | Infections per 100 persons<sup>a</sup> | Deaths per 100 persons<sup>a</sup> | Mild infections per 100 persons<sup>a</sup> | Severe and/or critical disease cases per 100 persons<sup>a</sup> | Day at peak incidence<sup>a</sup> |
| --- | --- | --- | --- | --- | --- | --- | --- | --- | --- |
| Country | Thousands | Years | Years | N (95% UI) | N (95% UI) | N (95% UI) | N (95% UI) | N (95% UI) | N (95% UI) |
| Afghanistan | 38,928,341 | 18.4 | 66.0 | 1.03 (0.94-1.55) | 2.5 (0.0-27.8) | 0.1 (0.0-0.6) | 2.5 (0.0-23.7) | 0.5 (0.0-4.1) | 700<sup>b</sup> (436.0-700<sup>b</sup>) |
| Bahrain | 1,701,583 | 32.5 | 77.7 | 1.65 (1.50-2.48) | 61.0 (49.0-66.7) | 0.9 (0.8-1.1) | 51.0 (42.1-57.4) | 8.7 (7.0-9.4) | 146.5 (141.0-154.0) |
| Egypt | 102,334,403 | 24.6 | 72.5 | 1.35 (1.22-2.01) | 37.0 (26.7-45.5) | 1.2 (0.9-1.3) | 31.0 (22.4-38.4) | 5.9 (4.3-7.1) | 285.0 (255.0-350.0) |
| Iran | 83,992,953 | 32.0 | 77.3 | 1.55 (1.47-2.41) | 53.0 (43.9-60.2) | 1.7 (1.4-1.8) | 44.5 (36.8-50.8) | 8.7 (7.1-9.4) | 195.0 (187.0-207.0) |
| Iraq | 40,222,503 | 21.0 | 71.1 | 1.15 (1.06-1.75) | 27.0 (14.3-36.2) | 0.7 (0.4-0.8) | 23.0 (12.1-30.8) | 4.1 (2.2-5.4) | 375.0 (320.0-628.0) |
| Jordan | 10,203,140 | 23.8 | 75.0 | 1.25 (1.17-1.93) | 37.0 (23.5-43.5) | 0.9 (0.6-1.0) | 29.0 (19.9-37.0) | 5.3 (3.6-6.5) | 265.0 (238.0-353.0) |
| Kuwait | 4,270,563 | 36.8 | 75.9 | 1.75 (1.59-2.61) | 61.0 (51.6-67.0) | 1.2 (1.1-1.4) | 51.5 (43.9-57.1) | 9.1 (7.8-9.9) | 143.5 (139.0-147.0) |
| Lebanon | 6,825,442 | 29.6 | 79.3 | 1.55 (1.43-2.34) | 51.0 (40.5-57.2) | 1.8 (1.5-1.9) | 41.5 (33.8-48.0) | 8.3 (6.7-9.2) | 175.0 (168.0-188.0) |
| Libya | 6,871,287 | 28.8 | 73.4 | 1.45 (1.34-2.20) | 45.0 (35.4-53.7) | 1.1 (0.9-1.2) | 39.0 (29.9-45.7) | 7.1 (5.4-8.0) | 196.5 (184.0-222.0) |
| Morocco | 36,910,558 | 29.5 | 77.4 | 1.55 (1.42-2.33) | 49.0 (39.7-56.2) | 1.7 (1.5-1.9) | 40.5 (33.1-47.1) | 8.3 (6.7-9.2) | 197.0 (188.0-213.0) |
| Oman | 5,106,622 | 30.6 | 78.6 | 1.55 (1.39-2.30) | 53.0 (41.9-61.4) | 0.7 (0.7-0.9) | 47.0 (36.1-53.0) | 7.4 (5.9-8.4) | 178.5 (170.0-199.0) |
| Pakistan | 220,892,331 | 22.8 | 67.8 | 1.25 (1.14-1.88) | 33.0 (21.8-41.3) | 0.9 (0.7-1.1) | 27.0 (18.3-35.0) | 5.0 (3.4-6.3) | 350.0 (303.0-479.0) |
| Palestine | 5,101,416 | 20.8 | 74.6 | 1.15 (1.04-1.71) | 25.0 (12.3-34.5) | 0.6 (0.3-0.8) | 19.0 (10.4-29.4) | 3.8 (1.9-5.1) | 345.0 (293.0-627.0) |
| Qatar | 2,881,060 | 32.3 | 80.7 | 1.65 (1.54-2.55) | 65.0 (53.6-71.5) | 0.8 (0.7-0.9) | 55.0 (46.3-61.8) | 8.9 (7.4-9.7) | 147.5 (141.0-153.0) |
| Saudi Arabia | 34,813,867 | 31.8 | 75.7 | 1.55 (1.42-2.35) | 53.0 (42.1-59.7) | 1.0 (0.9-1.1) | 45.0 (35.9-51.1) | 7.9 (6.2-8.7) | 193.5 (185.0-212.0) |
| Somalia | 15,893,219 | 16.7 | 58.3 | 0.98 (0.88-1.45) | 1.5 (0.0-22.8) | 0.0 (0.0-0.5) | 1.5 (0.0-19.4) | 0.3 (0.0-3.4) | 697.5 (501.0-700<sup>b</sup>) |
| Sudan | 43,849,269 | 19.7 | 66.1 | 1.15 (1.02-1.68) | 23.0 (1.7-32.9) | 0.7 (0.0-0.8) | 21.0 (1.5-27.9) | 3.7 (0.3-5.0) | 405.0 (352.0-700<sup>b</sup>) |
| Syria | 17,500,657 | 25.6 | 76.1 | 1.35 (1.24-2.05) | 39.0 (28.6-48.0) | 1.1 (0.8-1.2) | 33.0 (24.2-40.7) | 6.2 (4.5-7.3) | 242.5 (222.0-295.0) |
| Tunisia | 11,818,618 | 32.8 | 77.4 | 1.65 (1.53-2.50) | 55.0 (46.3-61.5) | 2.0 (1.8-2.2) | 45.5 (38.4-51.3) | 9.5 (7.8-10.2) | 162.5 (158.0-168.0) |
| UAE | 9,890,400 | 32.6 | 78.5 | 1.65 (1.53-2.54) | 63.0 (52.8-70.6) | 0.7 (0.6-0.8) | 55.0 (45.6-61.2) | 8.7 (7.2-9.5) | 161.0 (154.0-169.0) |
| Yemen | 29,825,968 | 20.2 | 66.4 | 1.05 (1.00-1.66) | 23.0 (0.4-32.8) | 0.6 (0.0-0.7) | 19.0 (0.4-28.0) | 3.4 (0.1-4.8) | 425.0 (357.0-700<sup>b</sup>) |
| **EMRO** | **729,834,200** | **28.8** | **75.7<sup>c</sup>** | **1.45 (0.98-1.75)<sup>c</sup>** | **45.0 (1.5-65.0)<sup>c</sup>** | **0.9 (0.0-2.0)<sup>c</sup>** | **39.0 (1.5-55.0)<sup>c</sup>** | **7.1 (0.3-9.5)<sup>c</sup>** | **197.0 (143.5-700<sup>b</sup>)<sup>c</sup>** |

N, number. Syria, Syrian Arab Republic. UAE, United Arab Emirates. UI, uncertainty interval.
<sup>a</sup>For each epidemiologic indicator, the table reports the most probable estimate and the 95% uncertainty interval. For a number of countries, $R_0$ was close to 1 in some of the uncertainty runs leading to an estimated time for peak incidence beyond the simulation duration of two years (delayed epidemic peak). For these countries, the upper limit of the uncertainty interval was set at 700 days, that is the end day of the simulation. Uncertainty runs in which $R_0$ was less than 1 led to no epidemic emergence, and thus were excluded from further analysis. Reported ranges for the epidemic indicators did not include these runs, except for the range of $R_0$ .
<sup>b</sup>The estimated time for peak incidence was beyond the simulation duration of two years. The upper limit of the uncertainty interval was set at 700 days, that is the end day of the simulation.
<sup>c</sup>Estimates are for the median (and range).

**Table S6:**
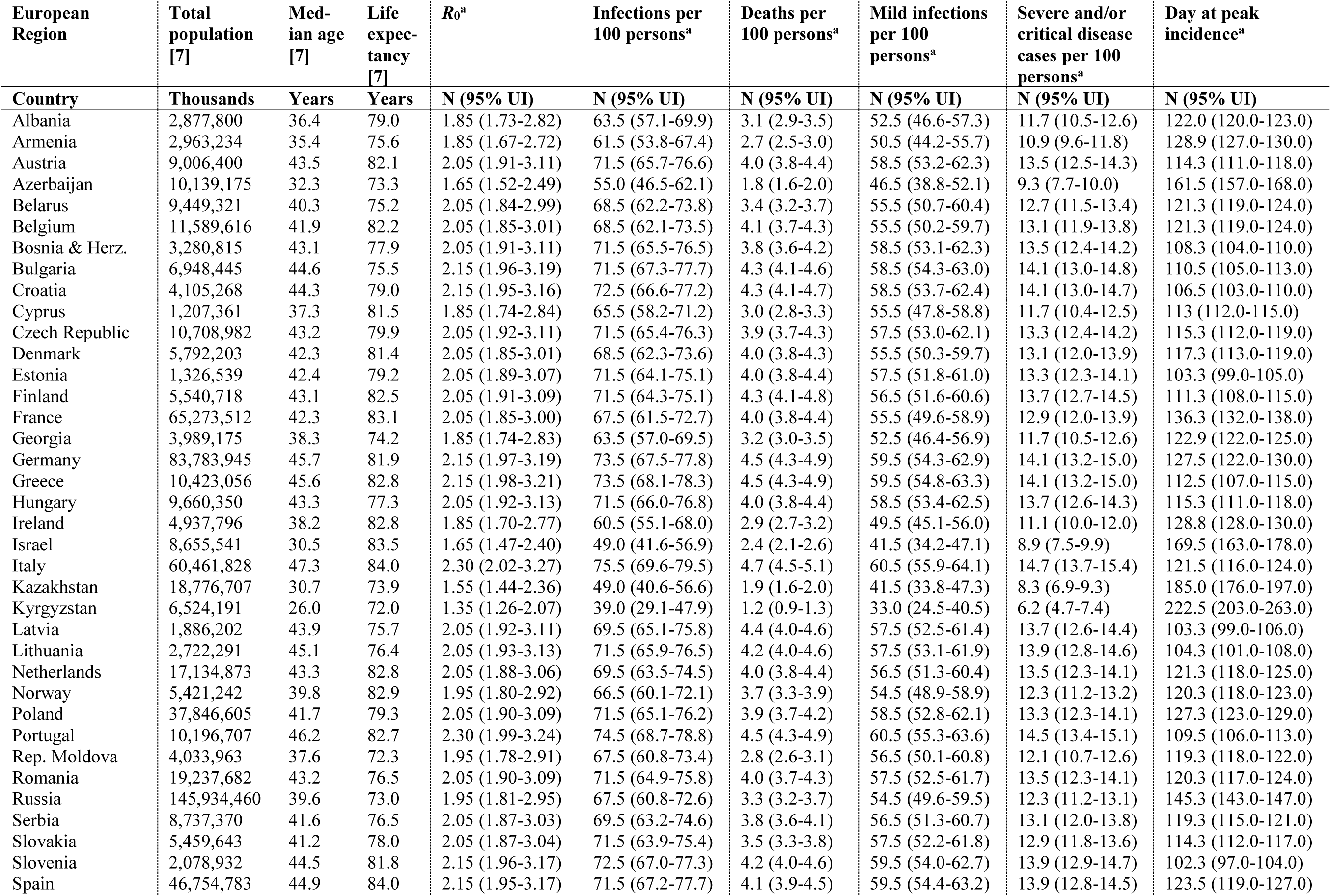
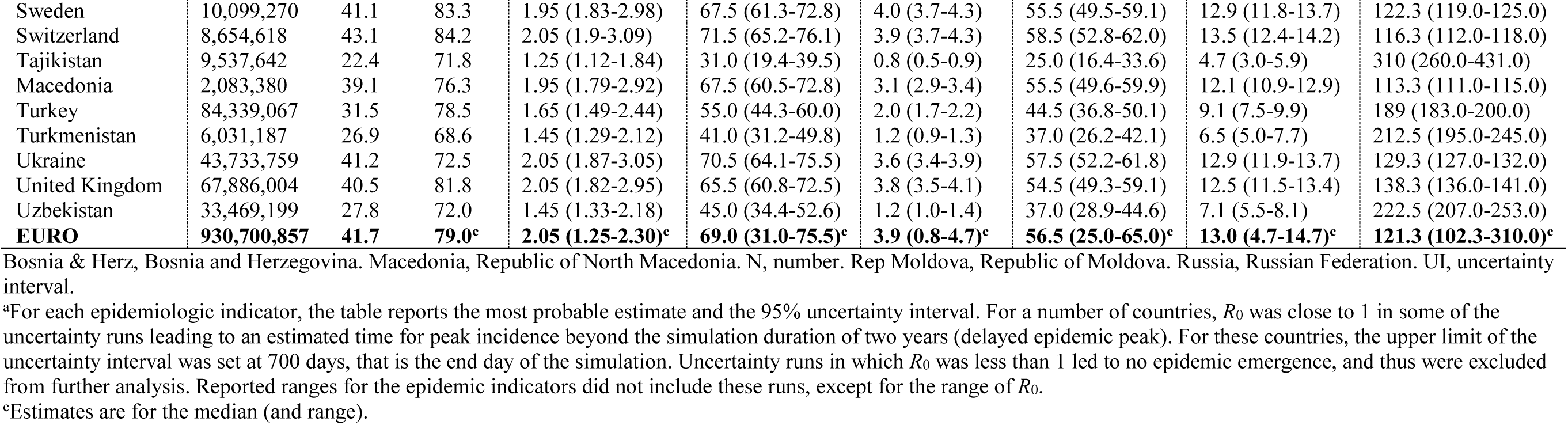
**Model estimates for key SARS-CoV-2 epidemiologic indicators for countries and territories with a population of at least one million [7] in the World Health Organization European Region (EURO)**.

| European Region | Total population [7] | Median age [7] | Life expectancy [7] | $R_0^a$ | Infections per 100 persons<sup>a</sup> | Deaths per 100 persons<sup>a</sup> | Mild infections per 100 persons<sup>a</sup> | Severe and/or critical disease cases per 100 persons<sup>a</sup> | Day at peak incidence<sup>a</sup> |
| --- | --- | --- | --- | --- | --- | --- | --- | --- | --- |
| Country | Thousands | Years | Years | N (95% UI) | N (95% UI) | N (95% UI) | N (95% UI) | N (95% UI) | N (95% UI) |
| Albania | 2,877,800 | 36.4 | 79.0 | 1.85 (1.73-2.82) | 63.5 (57.1-69.9) | 3.1 (2.9-3.5) | 52.5 (46.6-57.3) | 11.7 (10.5-12.6) | 122.0 (120.0-123.0) |
| Armenia | 2,963,234 | 35.4 | 75.6 | 1.85 (1.67-2.72) | 61.5 (53.8-67.4) | 2.7 (2.5-3.0) | 50.5 (44.2-55.7) | 10.9 (9.6-11.8) | 128.9 (127.0-130.0) |
| Austria | 9,006,400 | 43.5 | 82.1 | 2.05 (1.91-3.11) | 71.5 (65.7-76.6) | 4.0 (3.8-4.4) | 58.5 (53.2-62.3) | 13.5 (12.5-14.3) | 114.3 (111.0-118.0) |
| Azerbaijan | 10,139,175 | 32.3 | 73.3 | 1.65 (1.52-2.49) | 55.0 (46.5-62.1) | 1.8 (1.6-2.0) | 46.5 (38.8-52.1) | 9.3 (7.7-10.0) | 161.5 (157.0-168.0) |
| Belarus | 9,449,321 | 40.3 | 75.2 | 2.05 (1.84-2.99) | 68.5 (62.2-73.8) | 3.4 (3.2-3.7) | 55.5 (50.7-60.4) | 12.7 (11.5-13.4) | 121.3 (119.0-124.0) |
| Belgium | 11,589,616 | 41.9 | 82.2 | 2.05 (1.85-3.01) | 68.5 (62.1-73.5) | 4.1 (3.7-4.3) | 55.5 (50.2-59.7) | 13.1 (11.9-13.8) | 121.3 (119.0-124.0) |
| Bosnia & Herz. | 3,280,815 | 43.1 | 77.9 | 2.05 (1.91-3.11) | 71.5 (65.5-76.5) | 3.8 (3.6-4.2) | 58.5 (53.1-62.3) | 13.5 (12.4-14.2) | 108.3 (104.0-110.0) |
| Bulgaria | 6,948,445 | 44.6 | 75.5 | 2.15 (1.96-3.19) | 71.5 (67.3-77.7) | 4.3 (4.1-4.6) | 58.5 (54.3-63.0) | 14.1 (13.0-14.8) | 110.5 (105.0-113.0) |
| Croatia | 4,105,268 | 44.3 | 79.0 | 2.15 (1.95-3.16) | 72.5 (66.6-77.2) | 4.3 (4.1-4.7) | 58.5 (53.7-62.4) | 14.1 (13.0-14.7) | 106.5 (103.0-110.0) |
| Cyprus | 1,207,361 | 37.3 | 81.5 | 1.85 (1.74-2.84) | 65.5 (58.2-71.2) | 3.0 (2.8-3.3) | 55.5 (47.8-58.8) | 11.7 (10.4-12.5) | 113 (112.0-115.0) |
| Czech Republic | 10,708,982 | 43.2 | 79.9 | 2.05 (1.92-3.11) | 71.5 (65.4-76.3) | 3.9 (3.7-4.3) | 57.5 (53.0-62.1) | 13.3 (12.4-14.2) | 115.3 (112.0-119.0) |
| Denmark | 5,792,203 | 42.3 | 81.4 | 2.05 (1.85-3.01) | 68.5 (62.3-73.6) | 4.0 (3.8-4.3) | 55.5 (50.3-59.7) | 13.1 (12.0-13.9) | 117.3 (113.0-119.0) |
| Estonia | 1,326,539 | 42.4 | 79.2 | 2.05 (1.89-3.07) | 71.5 (64.1-75.1) | 4.0 (3.8-4.4) | 57.5 (51.8-61.0) | 13.3 (12.3-14.1) | 103.3 (99.0-105.0) |
| Finland | 5,540,718 | 43.1 | 82.5 | 2.05 (1.91-3.09) | 71.5 (64.3-75.1) | 4.3 (4.1-4.8) | 56.5 (51.6-60.6) | 13.7 (12.7-14.5) | 111.3 (108.0-115.0) |
| France | 65,273,512 | 42.3 | 83.1 | 2.05 (1.85-3.00) | 67.5 (61.5-72.7) | 4.0 (3.8-4.4) | 55.5 (49.6-58.9) | 12.9 (12.0-13.9) | 136.3 (132.0-138.0) |
| Georgia | 3,989,175 | 38.3 | 74.2 | 1.85 (1.74-2.83) | 63.5 (57.0-69.5) | 3.2 (3.0-3.5) | 52.5 (46.4-56.9) | 11.7 (10.5-12.6) | 122.9 (122.0-125.0) |
| Germany | 83,783,945 | 45.7 | 81.9 | 2.15 (1.97-3.19) | 73.5 (67.5-77.8) | 4.5 (4.3-4.9) | 59.5 (54.3-62.9) | 14.1 (13.2-15.0) | 127.5 (122.0-130.0) |
| Greece | 10,423,056 | 45.6 | 82.8 | 2.15 (1.98-3.21) | 73.5 (68.1-78.3) | 4.5 (4.3-4.9) | 59.5 (54.8-63.3) | 14.1 (13.2-15.0) | 112.5 (107.0-115.0) |
| Hungary | 9,660,350 | 43.3 | 77.3 | 2.05 (1.92-3.13) | 71.5 (66.0-76.8) | 4.0 (3.8-4.4) | 58.5 (53.4-62.5) | 13.7 (12.6-14.3) | 115.3 (111.0-118.0) |
| Ireland | 4,937,796 | 38.2 | 82.8 | 1.85 (1.70-2.77) | 60.5 (55.1-68.0) | 2.9 (2.7-3.2) | 49.5 (45.1-56.0) | 11.1 (10.0-12.0) | 128.8 (128.0-130.0) |
| Israel | 8,655,541 | 30.5 | 83.5 | 1.65 (1.47-2.40) | 49.0 (41.6-56.9) | 2.4 (2.1-2.6) | 41.5 (34.2-47.1) | 8.9 (7.5-9.9) | 169.5 (163.0-178.0) |
| Italy | 60,461,828 | 47.3 | 84.0 | 2.30 (2.02-3.27) | 75.5 (69.6-79.5) | 4.7 (4.5-5.1) | 60.5 (55.9-64.1) | 14.7 (13.7-15.4) | 121.5 (116.0-124.0) |
| Kazakhstan | 18,776,707 | 30.7 | 73.9 | 1.55 (1.44-2.36) | 49.0 (40.6-56.6) | 1.9 (1.6-2.0) | 41.5 (33.8-47.3) | 8.3 (6.9-9.3) | 185.0 (176.0-197.0) |
| Kyrgyzstan | 6,524,191 | 26.0 | 72.0 | 1.35 (1.26-2.07) | 39.0 (29.1-47.9) | 1.2 (0.9-1.3) | 33.0 (24.5-40.5) | 6.2 (4.7-7.4) | 222.5 (203.0-263.0) |
| Latvia | 1,886,202 | 43.9 | 75.7 | 2.05 (1.92-3.11) | 69.5 (65.1-75.8) | 4.4 (4.0-4.6) | 57.5 (52.5-61.4) | 13.7 (12.6-14.4) | 103.3 (99.0-106.0) |
| Lithuania | 2,722,291 | 45.1 | 76.4 | 2.05 (1.93-3.13) | 71.5 (65.9-76.5) | 4.2 (4.0-4.6) | 57.5 (53.1-61.9) | 13.9 (12.8-14.6) | 104.3 (101.0-108.0) |
| Netherlands | 17,134,873 | 43.3 | 82.8 | 2.05 (1.88-3.06) | 69.5 (63.5-74.5) | 4.0 (3.8-4.4) | 56.5 (51.3-60.4) | 13.5 (12.3-14.1) | 121.3 (118.0-125.0) |
| Norway | 5,421,242 | 39.8 | 82.9 | 1.95 (1.80-2.92) | 66.5 (60.1-72.1) | 3.7 (3.3-3.9) | 54.5 (48.9-58.9) | 12.3 (11.2-13.2) | 120.3 (118.0-123.0) |
| Poland | 37,846,605 | 41.7 | 79.3 | 2.05 (1.90-3.09) | 71.5 (65.1-76.2) | 3.9 (3.7-4.2) | 58.5 (52.8-62.1) | 13.3 (12.3-14.1) | 127.3 (123.0-129.0) |
| Portugal | 10,196,707 | 46.2 | 82.7 | 2.30 (1.99-3.24) | 74.5 (68.7-78.8) | 4.5 (4.3-4.9) | 60.5 (55.3-63.6) | 14.5 (13.4-15.1) | 109.5 (106.0-113.0) |
| Rep. Moldova | 4,033,963 | 37.6 | 72.3 | 1.95 (1.78-2.91) | 67.5 (60.8-73.4) | 2.8 (2.6-3.1) | 56.5 (50.1-60.8) | 12.1 (10.7-12.6) | 119.3 (118.0-122.0) |
| Romania | 19,237,682 | 43.2 | 76.5 | 2.05 (1.90-3.09) | 71.5 (64.9-75.8) | 4.0 (3.7-4.3) | 57.5 (52.5-61.7) | 13.5 (12.3-14.1) | 120.3 (117.0-124.0) |
| Russia | 145,934,460 | 39.6 | 73.0 | 1.95 (1.81-2.95) | 67.5 (60.8-72.6) | 3.3 (3.2-3.7) | 54.5 (49.6-59.5) | 12.3 (11.2-13.1) | 145.3 (143.0-147.0) |
| Serbia | 8,737,370 | 41.6 | 76.5 | 2.05 (1.87-3.03) | 69.5 (63.2-74.6) | 3.8 (3.6-4.1) | 56.5 (51.3-60.7) | 13.1 (12.0-13.8) | 119.3 (115.0-121.0) |
| Slovakia | 5,459,643 | 41.2 | 78.0 | 2.05 (1.87-3.04) | 71.5 (63.9-75.4) | 3.5 (3.3-3.8) | 57.5 (52.2-61.8) | 12.9 (11.8-13.6) | 114.3 (112.0-117.0) |
| Slovenia | 2,078,932 | 44.5 | 81.8 | 2.15 (1.96-3.17) | 72.5 (67.0-77.3) | 4.2 (4.0-4.6) | 59.5 (54.0-62.7) | 13.9 (12.9-14.7) | 102.3 (97.0-104.0) |
| Spain | 46,754,783 | 44.9 | 84.0 | 2.15 (1.95-3.17) | 71.5 (67.2-77.7) | 4.1 (3.9-4.5) | 59.5 (54.4-63.2) | 13.9 (12.8-14.5) | 123.5 (119.0-127.0) |
| Country | Thousands | Years | Years | N (95% UI) | N (95% UI) | N (95% UI) | N (95% UI) | N (95% UI) | N (95% UI) |
| Sweden | 10,099,270 | 41.1 | 83.3 | 1.95 (1.83-2.98) | 67.5 (61.3-72.8) | 4.0 (3.7-4.3) | 55.5 (49.5-59.1) | 12.9 (11.8-13.7) | 122.3 (119.0-125.0) |
| Switzerland | 8,654,618 | 43.1 | 84.2 | 2.05 (1.9-3.09) | 71.5 (65.2-76.1) | 3.9 (3.7-4.3) | 58.5 (52.8-62.0) | 13.5 (12.4-14.2) | 116.3 (112.0-118.0) |
| Tajikistan | 9,537,642 | 22.4 | 71.8 | 1.25 (1.12-1.84) | 31.0 (19.4-39.5) | 0.8 (0.5-0.9) | 25.0 (16.4-33.6) | 4.7 (3.0-5.9) | 310 (260.0-431.0) |
| Macedonia | 2,083,380 | 39.1 | 76.3 | 1.95 (1.79-2.92) | 67.5 (60.5-72.8) | 3.1 (2.9-3.4) | 55.5 (49.6-59.9) | 12.1 (10.9-12.9) | 113.3 (111.0-115.0) |
| Turkey | 84,339,067 | 31.5 | 78.5 | 1.65 (1.49-2.44) | 55.0 (44.3-60.0) | 2.0 (1.7-2.2) | 44.5 (36.8-50.1) | 9.1 (7.5-9.9) | 189 (183.0-200.0) |
| Turkmenistan | 6,031,187 | 26.9 | 68.6 | 1.45 (1.29-2.12) | 41.0 (31.2-49.8) | 1.2 (0.9-1.3) | 37.0 (26.2-42.1) | 6.5 (5.0-7.7) | 212.5 (195.0-245.0) |
| Ukraine | 43,733,759 | 41.2 | 72.5 | 2.05 (1.87-3.05) | 70.5 (64.1-75.5) | 3.6 (3.4-3.9) | 57.5 (52.2-61.8) | 12.9 (11.9-13.7) | 129.3 (127.0-132.0) |
| United Kingdom | 67,886,004 | 40.5 | 81.8 | 2.05 (1.82-2.95) | 65.5 (60.8-72.5) | 3.8 (3.5-4.1) | 54.5 (49.3-59.1) | 12.5 (11.5-13.4) | 138.3 (136.0-141.0) |
| Uzbekistan | 33,469,199 | 27.8 | 72.0 | 1.45 (1.33-2.18) | 45.0 (34.4-52.6) | 1.2 (1.0-1.4) | 37.0 (28.9-44.6) | 7.1 (5.5-8.1) | 222.5 (207.0-253.0) |
| **EURO** | **930,700,857** | **41.7** | **79.0<sup>c</sup>** | **2.05 (1.25-2.30)<sup>c</sup>** | **69.0 (31.0-75.5)<sup>c</sup>** | **3.9 (0.8-4.7)<sup>c</sup>** | **56.5 (25.0-65.0)<sup>c</sup>** | **13.0 (4.7-14.7)<sup>c</sup>** | **121.3 (102.3-310.0)<sup>c</sup>** |

Bosnia & Herz, Bosnia and Herzegovina. Macedonia, Republic of North Macedonia. N, number. Rep Moldova, Republic of Moldova. Russia, Russian Federation. UI, uncertainty interval.
<sup>a</sup>For each epidemiologic indicator, the table reports the most probable estimate and the 95% uncertainty interval. For a number of countries, $R_0$ was close to 1 in some of the uncertainty runs leading to an estimated time for peak incidence beyond the simulation duration of two years (delayed epidemic peak). For these countries, the upper limit of the uncertainty interval was set at 700 days, that is the end day of the simulation. Uncertainty runs in which $R_0$ was less than 1 led to no epidemic emergence, and thus were excluded from further analysis. Reported ranges for the epidemic indicators did not include these runs, except for the range of $R_0$ .
<sup>c</sup>Estimates are for the median (and range).

**Table S7:**
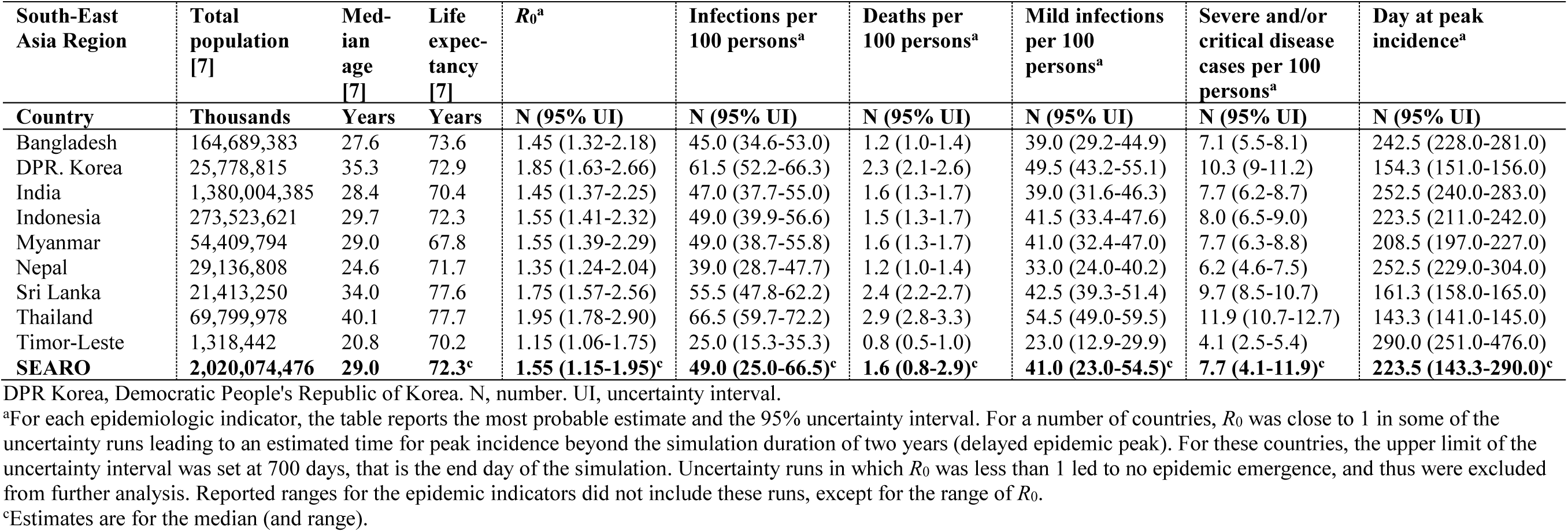
**Model estimates for key SARS-CoV-2 epidemiologic indicators for countries and territories with a population of at least one million [7] in the World Health Organization South-East Asia Region (SEARO)**.

**Table S8:**
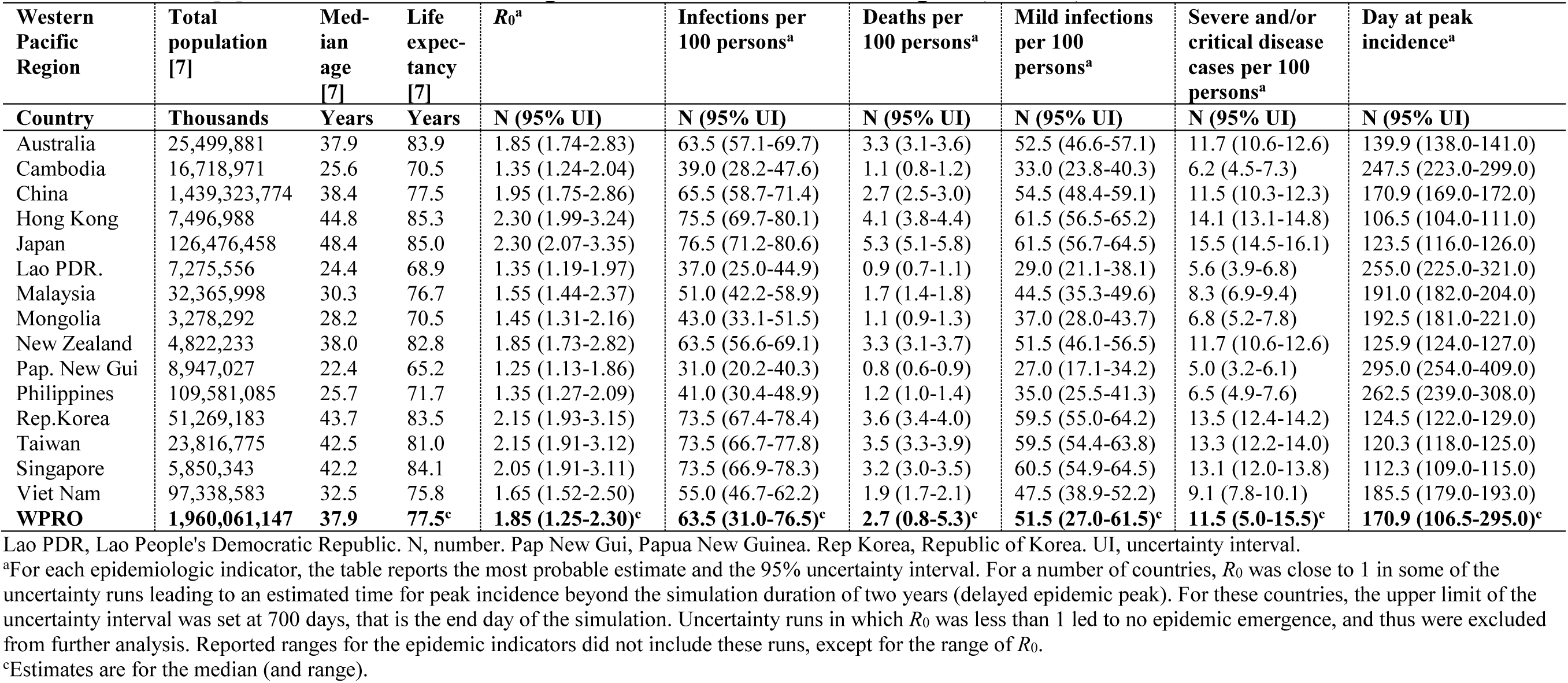
**Model estimates for key SARS-CoV-2 epidemiologic indicators for countries and territories with a population of at least one million [7] in the World Health Organization Western Pacific Region (WPRO)**.

**Figure S4:**
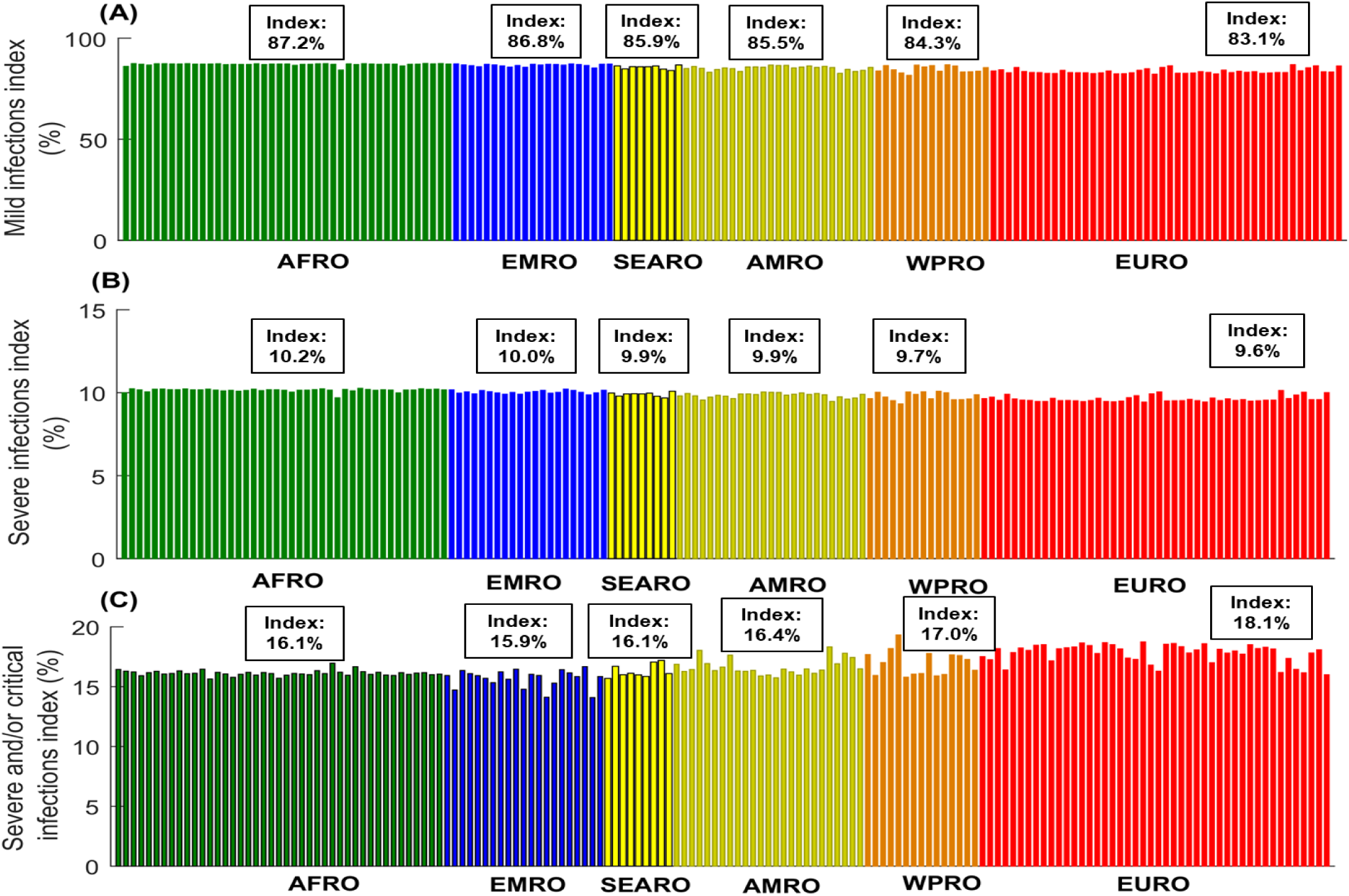
Estimates for the A) mild infections index, B) severe infections index, and C) severe and/or critical infections index in 159 countries and territories with a population of at least one million, across World Health Organization regions. These are the African Region (AFRO), Eastern Mediterranean Region (EMRO), South-East Asia Region (SEARO), Region of the Americas (AMRO), Western Pacific Region (WPRO), and European Region (EURO). The figure shows also the median index for each world region.

**Figure S5:**
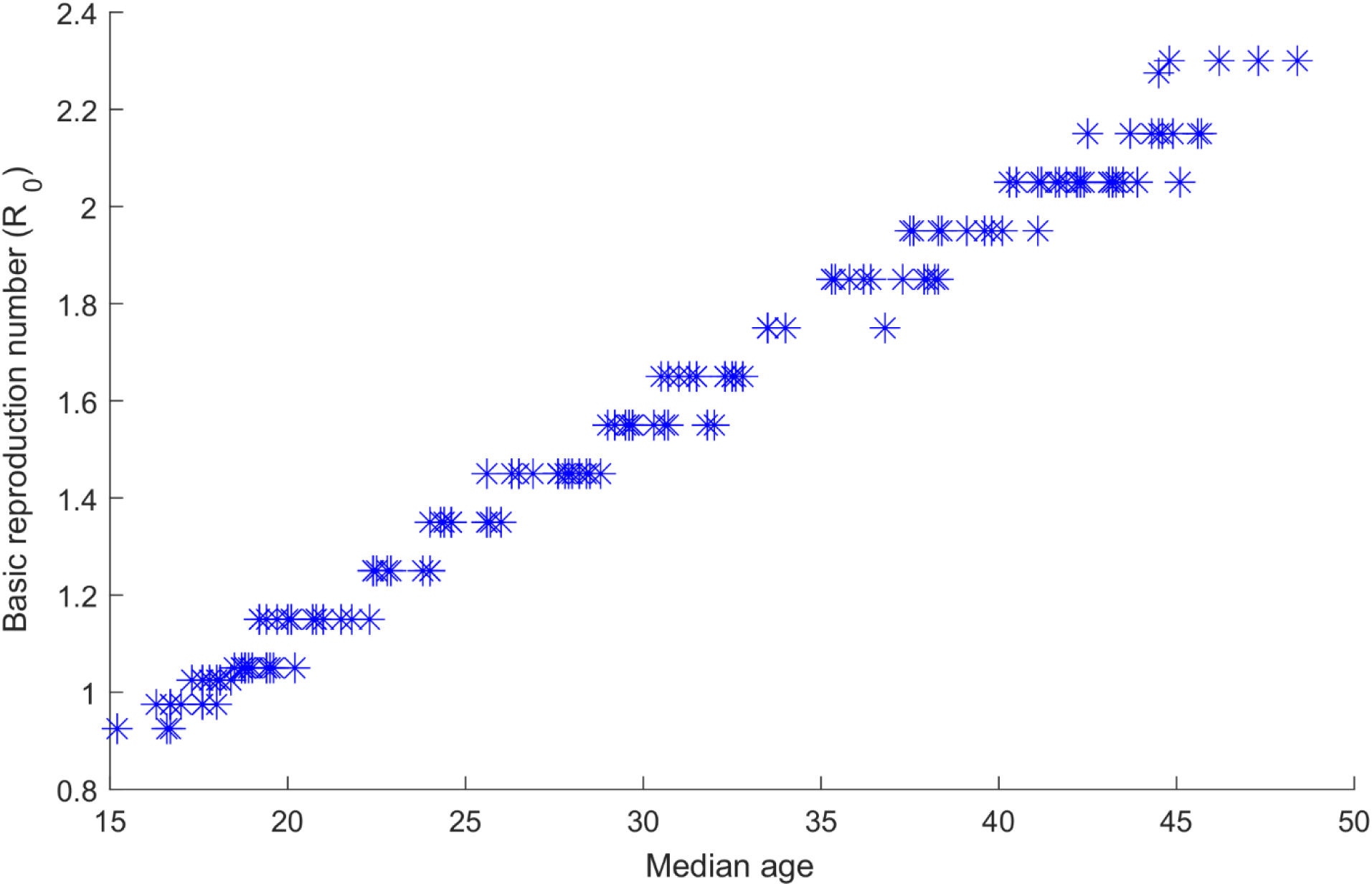
Impact of the variation in median age in any country on the basic reproduction number, *R*_0_. This figure was produced based on the estimates of all countries.

**Figure S6:**
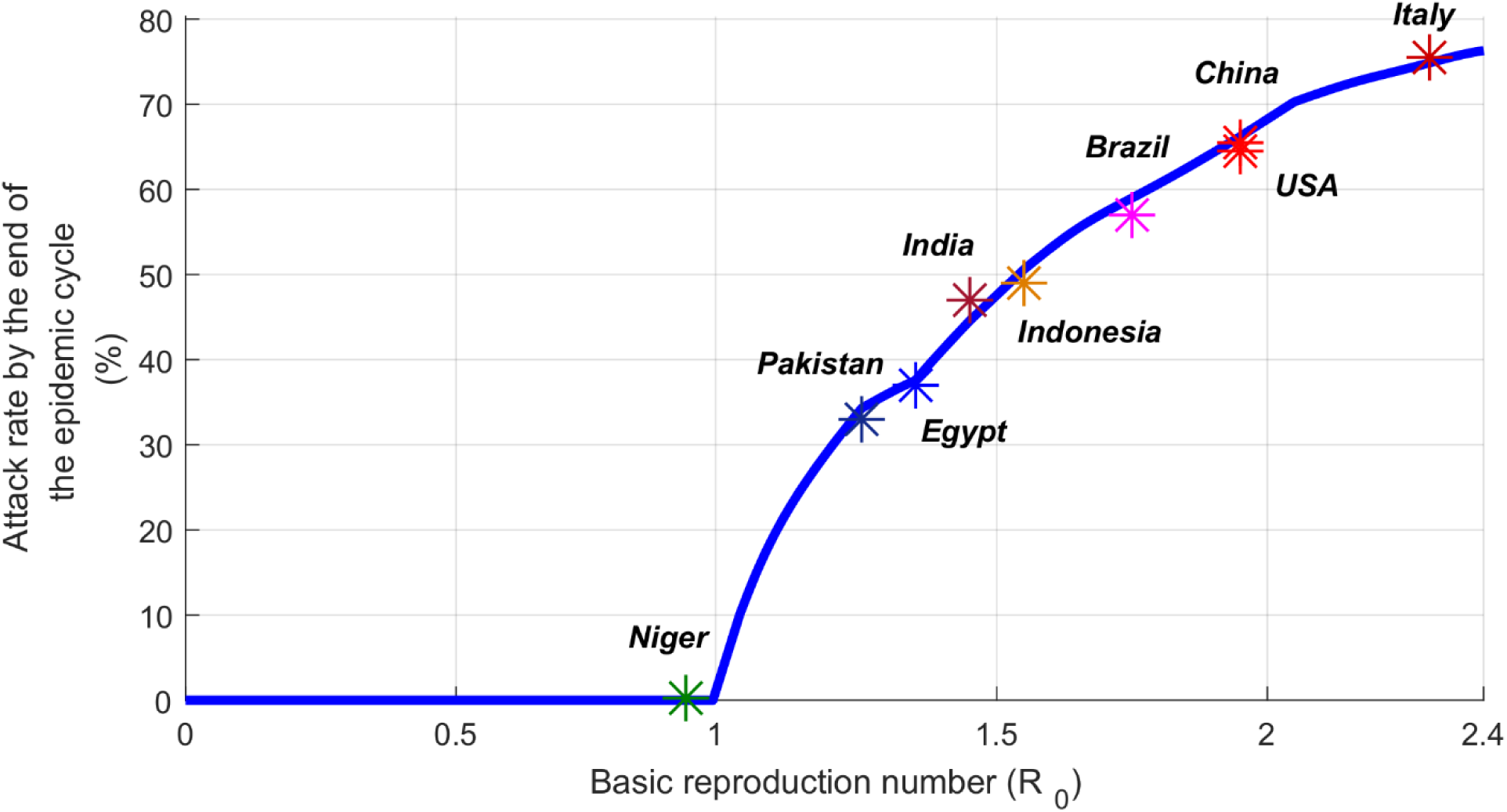
Impact of the variation in the basic reproduction number, *R*_0_, on the attack rate by the end of the epidemic cycle. This figure was produced by curve fitting the estimates of all countries, however, the figure highlights only the specific data points of the select countries presented in the main text.

**Figure S7:**
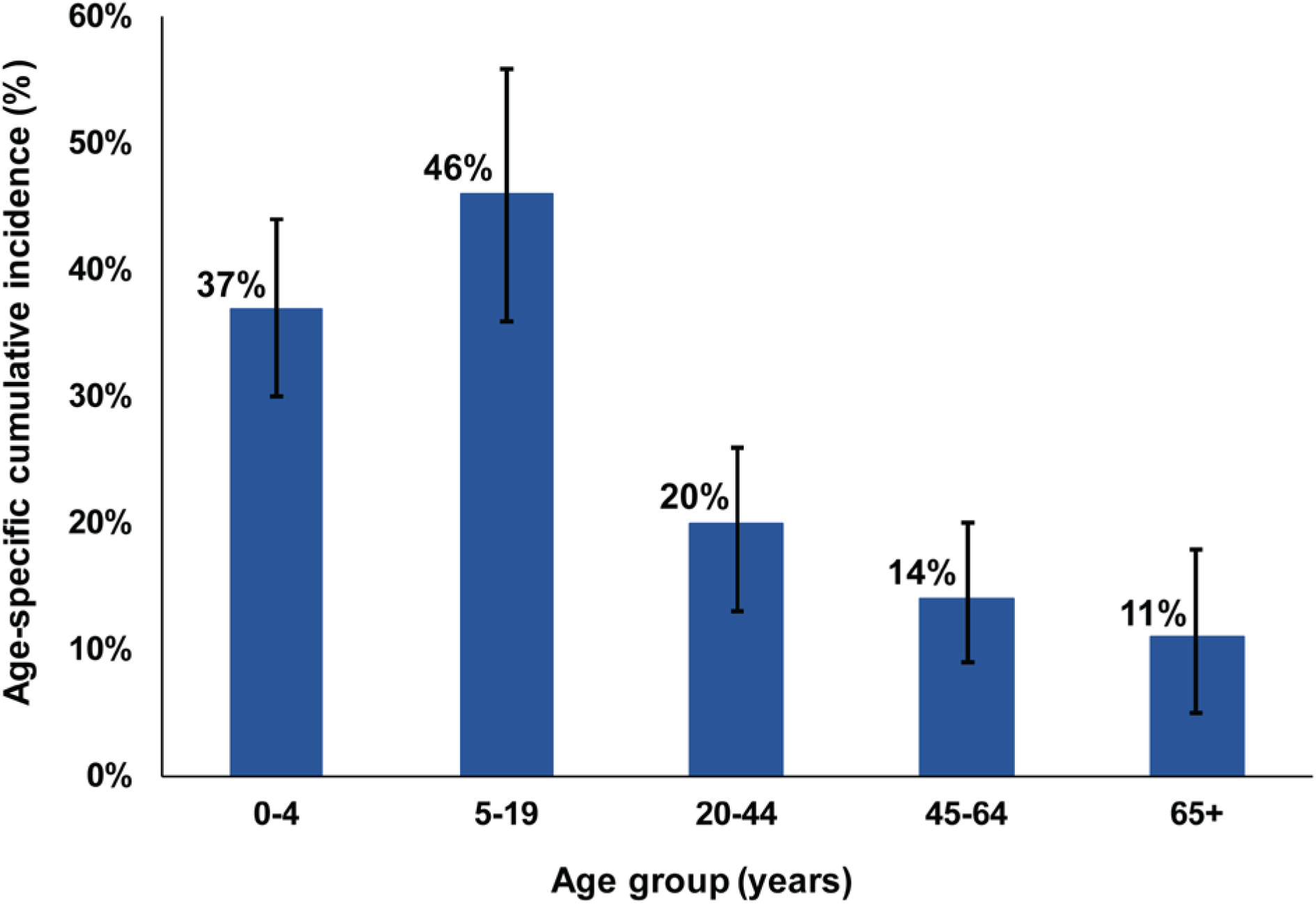
**Age-specific cumulative incidence of the 2009 influenza A (H1N1) pandemic (H1N1pdm) virus [13].**

**Figure S8:**
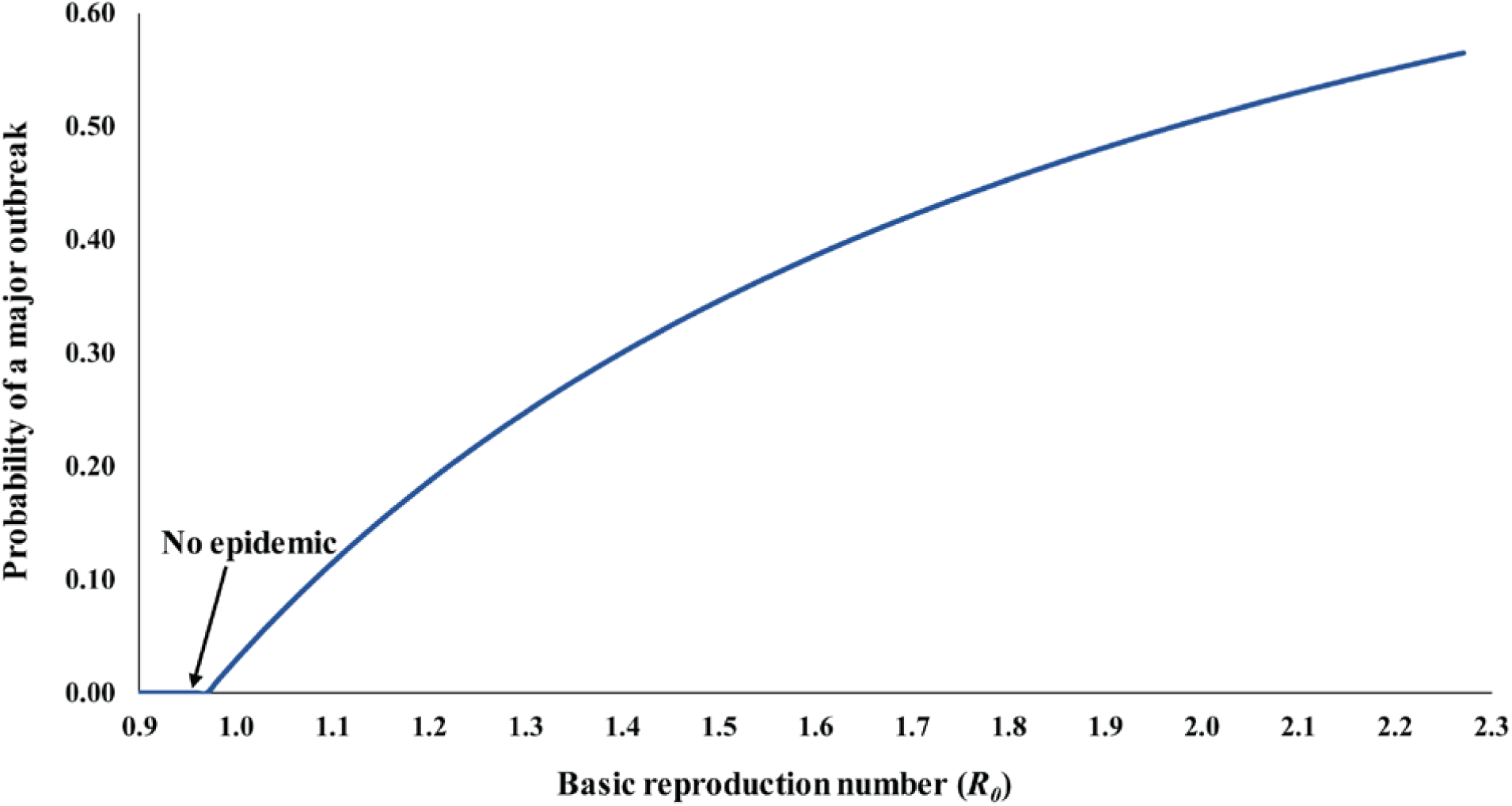
**Impact of the variation in the basic reproduction number, *R*_0_, on the probability of occurrence of a major SARS-CoV-2 outbreak upon introduction of one infection into the population.**

**Figure S9:**
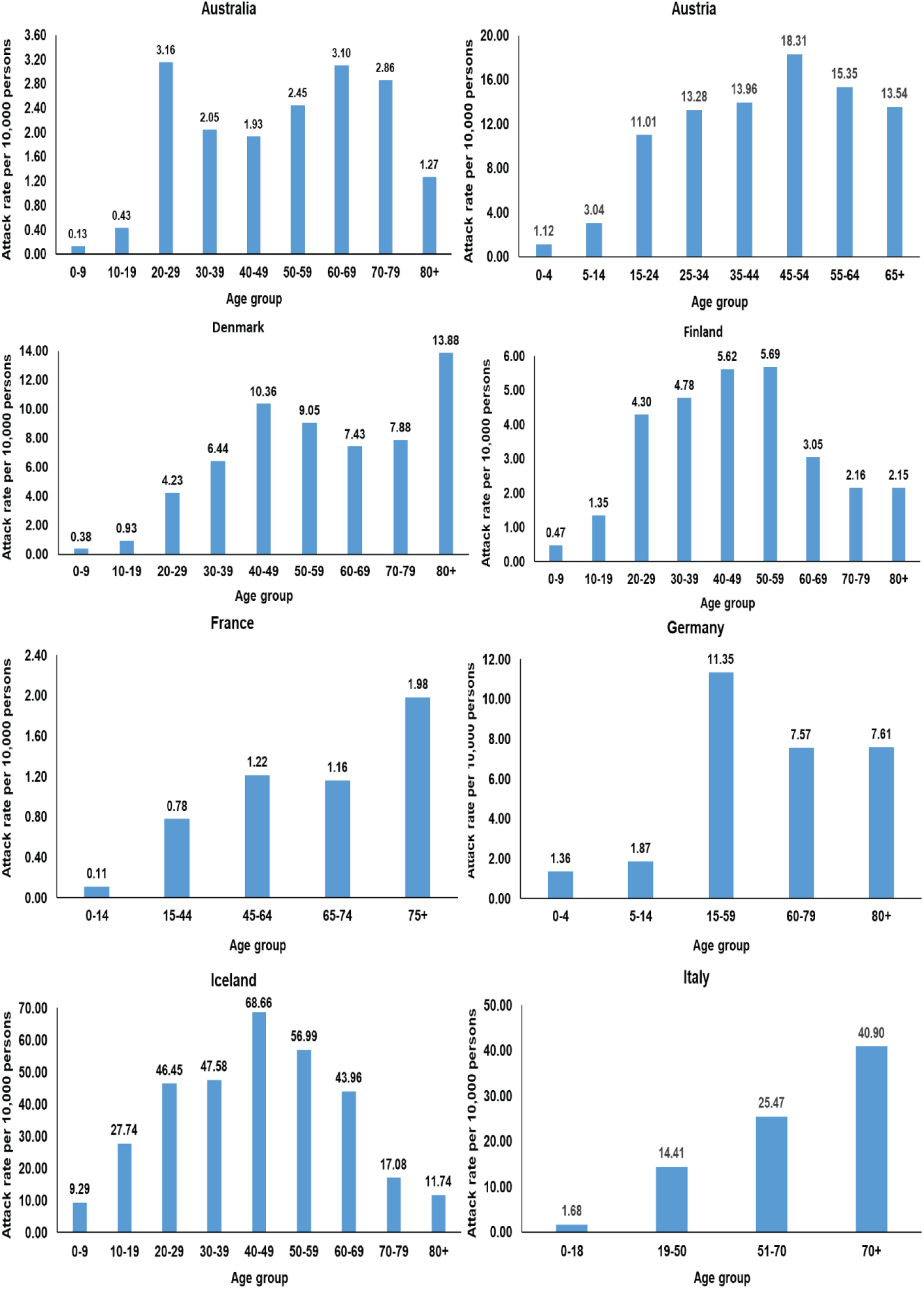
**Distribution of SARS-CoV-2 age-specific attack rate per 10**,**000 persons for Australia, Austria, Denmark, Finland, France, Germany, Iceland and Italy.**

**Figure S10:**
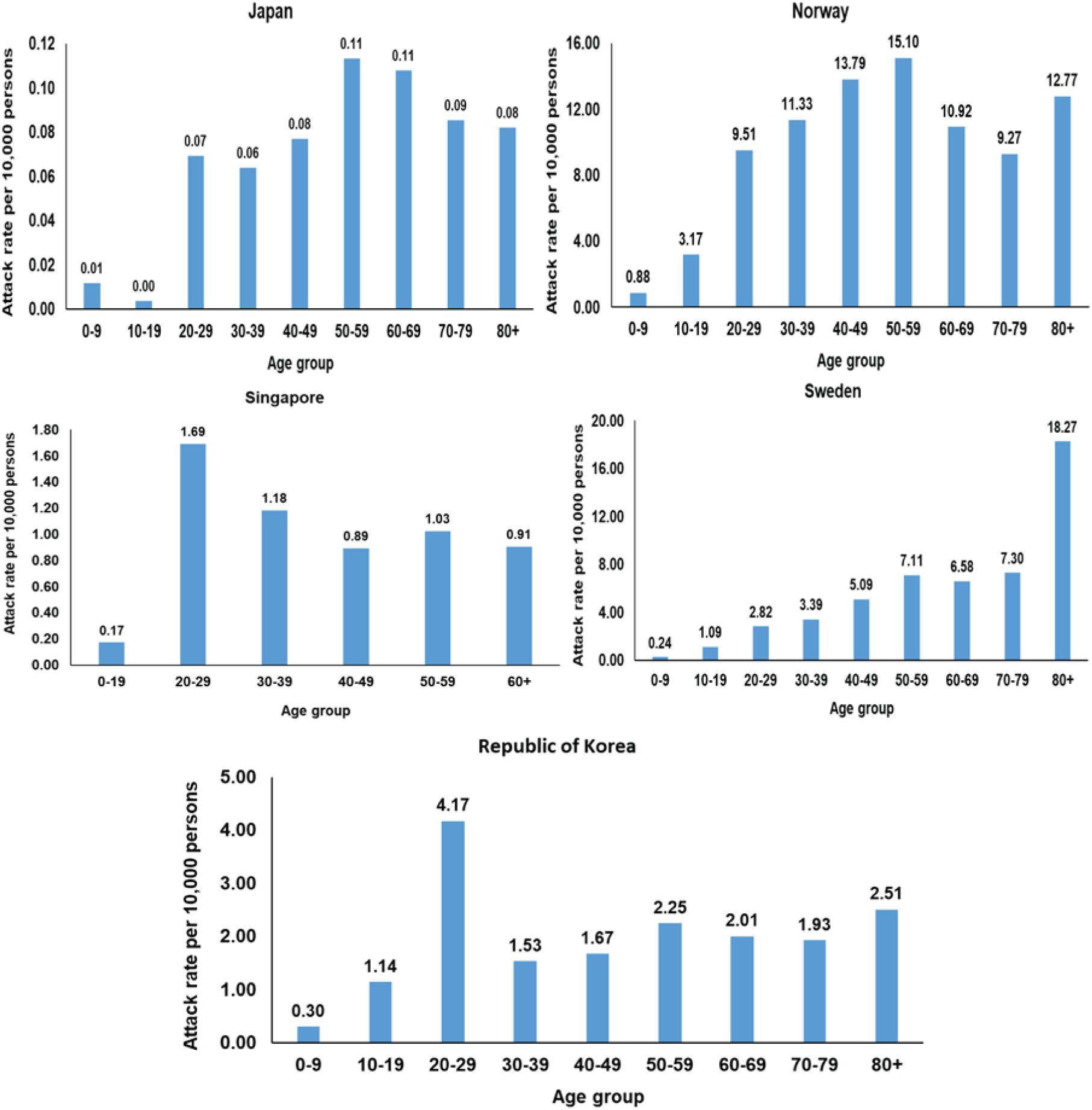
**Distribution of SARS-CoV-2 age-specific attack rate per 10**,**000 persons for Japan, Norway, Singapore, Sweden, and Republic of Korea.**

**Figure S11:**
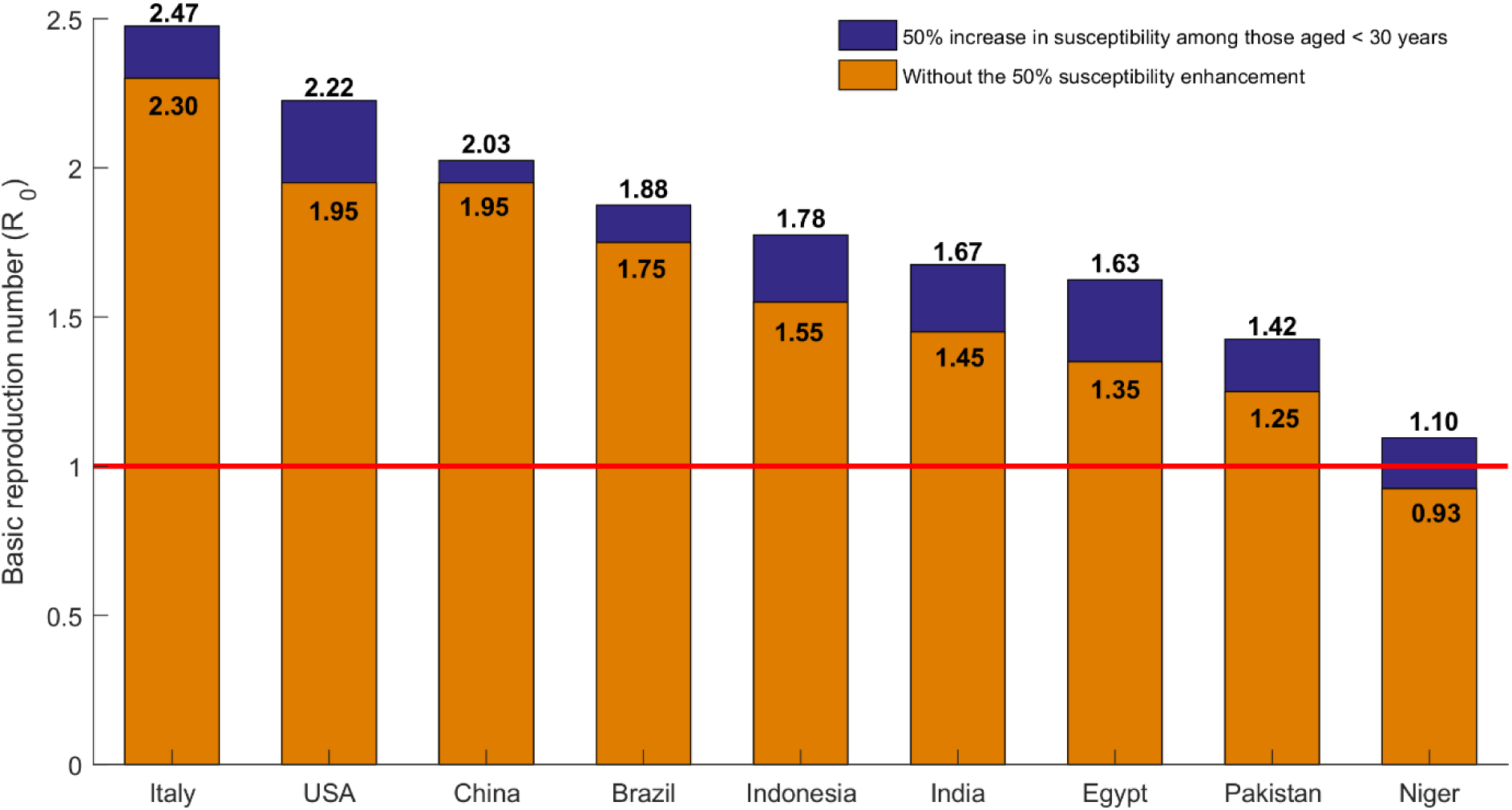
Sensitivity analysis assessing the impact of a 50% increase in the susceptibility to SARS-CoV-2 infection among those aged <30 years on our estimates for the basic reproduction number, *R*_0_, for the select countries presented in the main text. The figure also includes, for comparison, estimates of *R*_0_ without assuming the 50% susceptibility enhancement.

## References

1. World Health Organization (WHO), Naming the coronavirus disease (COVID-19) and the virus that causes it. Available from: https://www.who.int/emergencies/diseases/novel-coronavirus-2019/technical-guidance/naming-the-coronavirus-disease-(covid-2019)-and-the-virus-that-causes-it. Accessed on March 11, 2020. 2020.

2. COVID-19 Outbreak Live Update. Available from: https://www.worldometers.info/coronavirus/. Accessed on March 27, 2020. 2020.

3. Callaway, E., et al., The coronavirus pandemic in five powerful charts. Nature, 2020. 579(7800): p. 482–483.

4. Ayoub H.H., et al., Characterizing key attributes of the epidemiology of COVID-19 in China: Model-based estimations. under review.

5. Zhang, J., et al., Age profile of susceptibility, mixing, and social distancing shape the dynamics of the novel coronavirus disease 2019 outbreak in China. medRxiv, 2020: p. 2020.03.19.20039107.

6. Mizumoto, K., R. Omori, and H. Nishiura, Age specificity of cases and attack rate of novel coronavirus disease (COVID-19). medRxiv, 2020: p. 2020.03.09.20033142.

7. Davies, N.G., et al., Age-dependent effects in the transmission and control of COVID-19 epidemics. medRxiv, 2020: p. 2020.03.24.20043018.

8. World Health Organization, Report of the WHO-China Joint Mission on Coronavirus Disease 2019 (COVID-19). Available from :https://www.who.int/docs/default-source/coronaviruse/who-china-joint-mission-on-covid-19-final-report.pdf. Accessed on March 10, 2020. 2020.

9. Guan, W.J., et al., Clinical Characteristics of Coronavirus Disease 2019 in China. N Engl J Med, 2020.

10. Huang, C., et al., Clinical features of patients infected with 2019 novel coronavirus in Wuhan, China. Lancet, 2020. 395(10223): p. 497–506.

11. Novel Coronavirus Pneumonia Emergency Response Epidemiology, T., [The epidemiological characteristics of an outbreak of 2019 novel coronavirus diseases (COVID-19) in China]. Zhonghua Liu Xing Bing Xue Za Zhi, 2020. 41(2): p. 145–151.

12. Wu, Z. and J.M. McGoogan, Characteristics of and Important Lessons From the Coronavirus Disease 2019 (COVID-19) Outbreak in China: Summary of a Report of 72314 Cases From the Chinese Center for Disease Control and Prevention. JAMA, 2020.

13. Verity R., et al., Estimates of the severity of coronavirus disease 2019: a model-based analysis.The Lancet, 2020. 20: p. 30243–7.

14. United Nations Department of Economic and Social Affairs Population Dynamics, The 2019 Revision of World Population Prospects. Available from https://population.un.org/wpp/. Accessed on March 1st, 2020. 2020.

15. Li, R., et al., Substantial undocumented infection facilitates the rapid dissemination of novel coronavirus (SARS-CoV2). Science, 2020.

16. Lauer, S.A., et al., The Incubation Period of Coronavirus Disease 2019 (COVID-19) From Publicly Reported Confirmed Cases: Estimation and Application. Ann Intern Med, 2020.

17. Zou, L., et al., SARS-CoV-2 Viral Load in Upper Respiratory Specimens of Infected Patients. N Engl J Med, 2020.

18. Rothe, C., et al., Transmission of 2019-nCoV Infection from an Asymptomatic Contact in Germany. N Engl J Med, 2020. 382(10): p. 970–971.

19. Mckay, M.D., R.J. Beckman, and W.J. Conover, A Comparison of Three Methods for Selecting Values of Input Variables in the Analysis of Output from a Computer Code. Technometrics, 1979. 21(2): p. 239–245.

20. Sanchez, M.A. and S.M. Blower, Uncertainty and sensitivity analysis of the basic reproductive rate - Tuberculosis as an example. American Journal of Epidemiology, 1997. 145(12): p. 1127–1137.

21. MATLAB®, The Language of Technical Computing. The MathWorks, Inc. 2019.

22. StataCorp, Statistical Software: Release 16.1. College Station, TX: Stata Corporation, 2019.

23. Van Kerkhove, M.D., et al., Estimating age-specific cumulative incidence for the 2009 influenza pandemic: a meta-analysis of A(H1N1)pdm09 serological studies from 19 countries. Influenza Other Respir Viruses, 2013. 7(5): p. 872–86.

24. Whittle, P., The Outcome of a Stochastic Epidemic. Biometrika, 1955. 42(1-2): p. 116–122.

25. Allen, L.J.S. and G.E. Lahodny, Extinction thresholds in deterministic and stochastic epidemic models. Journal of Biological Dynamics, 2012. 6(2): p. 590–611.

26. Zhu, Y., et al., Children are unlikely to have been the primary source of household SARS-CoV-2 infections. medRxiv, 2020: p. 2020.03.26.20044826.

## References

1. Ayoub H.H., et al., Characterizing key attributes of the epidemiology of COVID-19 in China: Model-based estimations. under review.

2. Guan, W.J., et al., Clinical Characteristics of Coronavirus Disease 2019 in China. N Engl J Med, 2020.

3. Huang, C., et al., Clinical features of patients infected with 2019 novel coronavirus in Wuhan, China. Lancet, 2020. 395(10223): p. 497–506.

4. World Health Organization, Report of the WHO-China Joint Mission on Coronavirus Disease 2019 (COVID-19). Available from :https://www.who.int/docs/default-source/coronaviruse/who-china-joint-mission-on-covid-19-final-report.pdf. Accessed on March 10, 2020. 2020.

5. Novel Coronavirus Pneumonia Emergency Response Epidemiology, T., [The epidemiological characteristics of an outbreak of 2019 novel coronavirus diseases (COVID-19) in China]. Zhonghua Liu Xing Bing Xue Za Zhi, 2020. 41(2): p. 145–151.

6. Wu, Z. and J.M. McGoogan, Characteristics of and Important Lessons From the Coronavirus Disease 2019 (COVID-19) Outbreak in China: Summary of a Report of 72314 Cases From the Chinese Center for Disease Control and Prevention. JAMA, 2020.

7. United Nations Department of Economic and Social Affairs Population Dynamics, The 2019 Revision of World Population Prospects. Available from https://population.un.org/wpp/. Accessed on March 1st, 2020. 2020.

8. Li, R., et al., Substantial undocumented infection facilitates the rapid dissemination of novel coronavirus (SARS-CoV2). Science, 2020.

9. Lauer, S.A., et al., The Incubation Period of Coronavirus Disease 2019 (COVID-19) From Publicly Reported Confirmed Cases: Estimation and Application. Ann Intern Med, 2020.

10. Zou, L., et al., SARS-CoV-2 Viral Load in Upper Respiratory Specimens of Infected Patients. N Engl J Med, 2020.

11. Rothe, C., et al., Transmission of 2019-nCoV Infection from an Asymptomatic Contact in Germany. N Engl J Med, 2020. 382(10): p. 970–971.

12. Heffernan, J.M., R.J. Smith, and L.M. Wahl, Perspectives on the basic reproductive ratio. Journal of the Royal Society Interface, 2005. 2(4): p. 281–293.

13. Van Kerkhove, M.D., et al., Estimating age-specific cumulative incidence for the 2009 influenza pandemic: a meta-analysis of A(H1N1)pdm09 serological studies from 19 countries. Influenza Other Respir Viruses, 2013. 7(5): p. 872–86.

